# Could global warming cause a range expansion or shift of Lyme disease in the U.S. state of Maryland? A mathematical modeling approach

**DOI:** 10.1101/2025.08.31.25334788

**Authors:** Salihu S. Musa, Abba B. Gumel

## Abstract

Lyme disease, transmitted by ticks, is endemic in several regions of the United States (including the Northeast), and the lifecycle of ticks is significantly affected by changes in local climatic variables. In this study, we modeled the dynamics of Lyme disease across the U.S. state of Maryland. We used a mechanistic model, calibrated using case and temperature data, to assess the impact of temperature fluctuations on the geospatial distribution and burden of Lyme disease across Maryland. Our results demonstrate that tick activity and Lyme disease intensity peak when temperature reaches 17.0°C—20.5°C. We estimate that moderate projected global warming will cause a range expansion of Lyme disease, increasing burden in Central Maryland and extending risk into Western counties, while reducing the disease burden in Southern and most Eastern counties. High projected warming will cause a westward shift, with new Lyme disease hotspots emerging in Western counties, and reduction of burden in Central, Southern and Eastern regions. Maryland will experience reductions in overall Lyme disease burden under both projected global warming scenarios (with more reductions under the high warming scenario). Disease elimination is feasible using a hybrid strategy, which combines rodents baiting, habitat clearance, and personal protection against tick bites, with moderate coverages.

## 1 Introduction

Lyme disease, caused by the spirochete bacterium *Borrelia burgdorferi* (*B. burgdorferi*) and transmitted primarily by infected ticks, is the most prevalent vector-borne disease in the United States [1, 2]. Although data from the United States Centers for Disease Control and Prevention (CDC) show there are approximately 30,000 laboratory-confirmed Lyme disease cases in the United States (U.S.) annually, substantial under-reporting of Lyme disease cases suggests that the true burden is significantly higher [1–3]. Specifically, data from insurance companies estimates that approximately 476,000 individuals may be diagnosed and treated for Lyme disease in the U.S. annually (this figure likely includes suspected, not confirmed, cases of Lyme disease reported in the insurance company data [2]). The economic burden of Lyme disease is substantial, with direct medical costs estimated to range from $712 to $1,300 *per* patient *per* year [4–6]. Other recent estimates suggest that the cost *per* patient *per* year is $3,000 [2, 7], resulting in an estimated annual healthcare expenditure for Lyme disease to be in excess of $1 billion in the U.S. [2, 4, 6]. However, these estimates primarily capture direct medical costs only, and the overall broader economic impact of the disease is much higher when accounting for associated lost economic productivity and long-term complications, such as Lyme arthritis and post-treatment Lyme disease syndrome [8, 9]. Furthermore, the increasing incidence of Lyme disease in endemic areas imposes a substantial additional economic burden on public health systems, necessitating increased investment in surveillance, vector control efforts, and community education initiatives [3, 10]. Since its identification in Lyme, Connecticut in the 1970s (following an outbreak of arthritis cases among children [11–13]), Lyme disease has become endemic in several regions within the U.S., notably the Northeast, Upper-Midwest, Mid-Atlantic, and the Pacific coast, where environmental conditions (temperature, humidity, and host availability) sustain the tick-host-pathogen cycle [14, 15]. The State of Maryland is one of the ten states most impacted by Lyme disease in the U.S. (alongside Pennsylvania, New Jersey, New York, Connecticut, and other states [2]). It records an average of 2,000-3,000 reported Lyme disease cases annually [2, 16], and the estimated direct medical cost for treating Lyme disease is around $2,000 *per* patient *per* year, resulting in total annual healthcare expenditure of around $4 to 6 million based on national estimates [2, 7] (this cost rises to $40−60 million if the estimate from insurance companies is used) [2].

Lyme disease is primarily transmitted in the U.S. by two tick species: the blacklegged tick (*Ixodes scapularis* or *I. scapularis*) in the Northeast, Upper-Midwest, and Mid-Atlantic regions, and the western blacklegged tick (*Ixodes pacificus*) along the Pacific Coast [17]. The white-footed mouse (*Peromyscus leucopus* or *P. leucopus*) serves as the most competent and main reservoir host of *B. burgdorferi* in North America, maintaining *B. burgdorferi* without exhibiting symptoms, while the white-tailed deer supports tick reproduction (by serving as a source of blood meal for ticks, and a sustainer for ticks population) but do not directly contribute to pathogen transmission (i.e., white-tailed deer does not acquire or transmit Lyme disease) [13, 18]. Other rodents, such as the Norway rat (*Rattus norvegicus*), edible dormouse (*Glis glis*), and various *Apodemus* species, along with ground-dwelling birds like the American robin (*Turdus migratorius*), also act as reservoir hosts for Lyme disease [19, 20]. Humans (who are dead-end hosts, since they do not sustain bacteremia at levels sufficient for human-to-tick transmission [21, 22]) are also reservoir hosts for Lyme disease [2, 19]. The hosts primarily acquire Lyme disease from infected nymphal ticks, which are the main vectors for Lyme disease transmission (due to their small size and prolonged feeding, which facilitate undetected pathogen transfer to hosts) [1, 3, 10, 23, 24]. Although adult female ticks (both *I. scapularis* and *I. pacificus* species) can also transmit *B. burgdorferi*, their role in such transmission is quite limited (i.e., nymphs are the main transmitters of Lyme disease, in comparison to adult female ticks). Adult male ticks do not transmit Lyme disease, and they bite only briefly to obtain blood for mating and do not remain attached to the host long enough for transmission to occur [21, 25]. Larvae, which hatch uninfected, acquire *B. burgdorferi* by feeding on infected small mammals, such as the white-footed mouse [18, 21, 25] (the life cycles of *I. scapularis* and *B. burgdorferi* are depicted in Figure 1 [25]).

**Figure 1:**
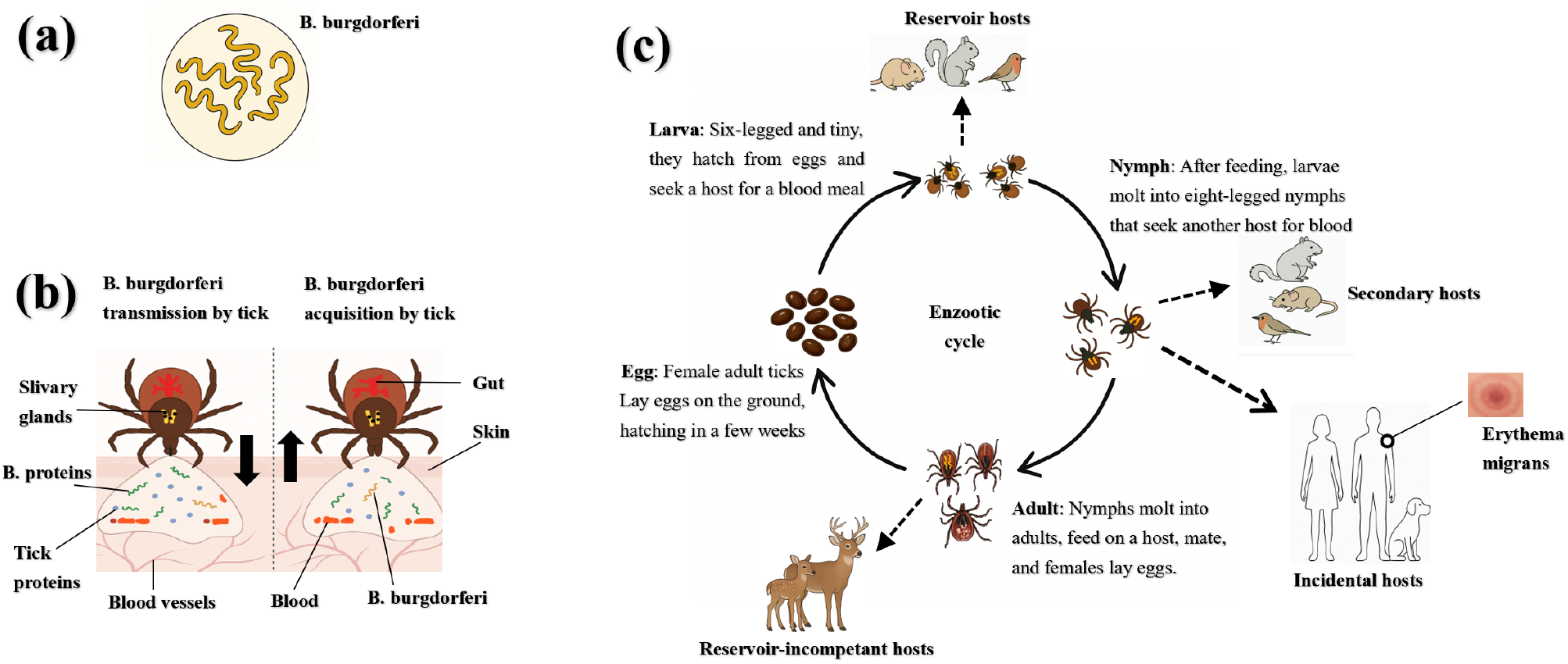
Lifecycles of *B. burgdorferi* and its transmission dynamics involving tick vectors and host populations. (**a**) Morphology (shape and structural characteristics) of *B. burgdorferi*, the spirochaete bacterium responsible for Lyme disease; (**b**) acquisition and transmission of *B. burgdorferi* via bite of blacklegged ticks (*I. scapularis*). During feeding on infected reservoir hosts (primarily white-footed mice), larval or nymphal ticks acquire the pathogen with the blood meal (right panel of (**b**)). Tick salivary proteins such as SALP25D reduce local inflammation, enhancing spirochaete survival at the bite site. Within the tick, the spirochaetes attach to the midgut epithelium via OspA and persist through molting. Upon the next blood meal, environmental cues activate expression of OspC and other proteins that facilitate migration to the salivary glands. Transmission to a new vertebrate host occurs via tick saliva (left panel of (**b**)), aided by salivary factors (e.g., SALP15, ISAC, TSLPI) that suppress immune responses and promote infection; (**c**) The enzootic cycle of *B. burgdorferi*, involving four developmental stages of *I. scapularis*—eggs, larva, nymph, and adult—each requiring a single blood meal (except eggs, which hatches to become larvae). Spirochaetes are acquired transstadially when larvae feed on infected small mammals or birds and are retained through molting. Nymphs, feeding on similar hosts, perpetuate the transmission cycle. Although adult ticks primarily feed on larger mammals like deer—which do not transmit the pathogen—they are essential for tick reproduction. Humans and domestic animals such as dogs may be bitten, but do not contribute to the maintenance of the bacterium in nature [12, 13]. Nymphs are the principal stage responsible for transmission to humans. All stages of the tick life cycle are influenced by ambient temperature.

Susceptible *I. scapularis* ticks (larvae, nymphs, or adult females) acquire *B. burgdorferi* infection following attachment to, and biting (for a blood meal), an infected competent host (human or mouse). There are two main mechanisms for infection of the tick, namely systemic (pathogen spreads via host bloodstream) and co-feeding-based (direct vector-to-vector transfer at feeding site) transmission [26, 27]. Upon infection, the spirochetes of the *B. burgdorferi* bacteria initially reside in the midgut of the tick. Before the infected tick can transmit infection, the *B. burgdorferi* bacteria require a reactivation period of 24–48 hours, during which they migrate from the midgut to the salivary glands, facilitating pathogen transmission to the host [11, 13]. Transmission risk increases with prolonged tick attachment to the host, as spirochetes enter the host dermis *via* the salivary duct, initiating infection [28]. Once inside the host (mice or humans), *B. burgdorferi* circulates through the bloodstream, evading immune responses by utilizing antigenic variation and immune-modulatory mechanisms [21, 22]. The bacterium subsequently establishes persistent infections in various tissues of the host, including the skin, joints, nervous system, and heart [11, 13]. In humans, early-stage Lyme disease typically manifests as *erythema migrans* (a distinctive skin rash, also known as *bull’s-eye rash*), fever, headache, and fatigue [1, 5, 11]. If untreated, the infection can progress, leading to Lyme arthritis, *neuroborreliosis*, and Lyme carditis, affecting the joints, central nervous system, and myocardium [1, 5, 11] (deaths due to Lyme disease are extremely rare [2]). Although there is currently no licensed human vaccine for Lyme disease, experimental oral vaccines targeting reservoir hosts such as white-footed mice have shown promise in reducing transmission of *B. burgdorferi* [29]. Furthermore, a multivalent human vaccine candidate (VLA15), developed by Valneva and Pfizer, is in Phase 3 clinical trials as of 2024, targeting six serotypes of the outer surface protein A (OspA) of *B. burgdorferi* [30, 31].

The disease has expanded its geographic range and burden in the U.S., over the past few decades, due to abiotic factors, such as climate change, ecological shifts, and anthropogenic landscape modifications [1–3, 10, 28, 32, 33], and biotic factors, such as host density, pathogen adaptation, and changes in vector population dynamics [34, 35]. Specifically, empirical studies have shown that rising temperatures, increased humidity, and habitat fragmentation have facilitated the northward and altitudinal expansion of *I. scapularis*, in addition to prolonging tick activity seasons and increasing human exposure risk in previously low-incidence regions [28, 36–39]. These changes in local climatic conditions also further increase Lyme disease risk by expanding the tick’s habitats, increasing vector densities, and extending seasonal transmission periods [33, 37]. This expansion of Lyme disease risk has heightened public health concerns, particularly in regions or jurisdictions within the United States with historically low (or even no) incidence of Lyme disease [2]. The reported incidence rates of Lyme disease in the U.S. have tripled over the last three decades, with the disease now endemic in at least 15 states, predominantly in the Northeast, Midwest, and Mid-Atlantic regions [2, 10]. Ecological and environmental (climatic) trends indicate a gradual shift in high-risk regions or areas, with tick populations establishing in northern latitudes and higher elevations that were once unsuitable habitats for their survival [39, 40]. These changes highlight the urgent need for realistic mathematical modeling to assess and predict the current and future trajectory of Lyme disease in regions that are currently at risk or potentially at risk for Lyme disease. In the State of Maryland, Lyme disease cases have risen steadily over the past two decades, with most of its 24 counties experiencing increasing incidence (with Montgomery County being the key hot-spot for Lyme disease transmission in the state [41]). The state’s diverse landscape—including coastal plains, agricultural zones, and densely forested regions—fosters complex tick-host dynamics that sustain Lyme disease transmission. Central and Western Maryland, with their extensive woodlands and high deer populations, have also become major transmission zones, reporting some of the highest infection rates in the mid-Atlantic region [9, 10]. Climate projections indicate that continued warming will increase tick habitat expansion, alter seasonal activity patterns, and exacerbate the Lyme disease burden in the state [33, 39]. These trends necessitate climate-adaptive vector management strategies to mitigate disease risk effectively.

Numerous mathematical models have been developed and used to investigate the ecological and epidemiological mechanisms governing the tick-host-pathogen lifecycle, particularly Lyme disease transmission [23, 42–61]. These models are of various types, including deterministic (compartmental) [45, 47, 49], stochastic [55, 56], statistical [57, 58], agent-based [39, 59, 60], and network [61] models, and were used to assess numerous pertinent aspects of the disease transmission dynamics and control, such as assessing the impact of climate change [37, 40, 44], biodiversity [15, 18, 62], land-use patterns [9, 63], and habitat fragmentation [49, 64] on tick population dynamics and Lyme disease epidemiology [1, 3, 23]. For instance, Ogden *et al*. [44] used a climate-driven deterministic model for the population dynamics of *I. scapularis* in Canada to show that rising temperatures accelerate tick development, extend host-seeking periods, and facilitate range expansion. This study was further extended by Ogden *et al*. [65] to predict northward shifts in *I. scapularis* populations in Canada. Wallace *et al*. [66] also used a mechanistic model to confirm a strong correlation between mean annual temperature and the establishment of *I. scapularis* in new regions, while Monaghan *et al*. [37] applied a statistical model to demonstrate that rising temperatures prolong seasonal tick activity, increasing transmission risk. Similarly, Wu *et al*. [46] estimated the basic reproduction number of *I. scapularis* in Canada (using the model in Ogden *et al*. [44]), showing that warming conditions facilitate the geographic expansion of Lyme disease risk, particularly in previously unsuitable northern regions of Canada.

Beyond climate, several modeling studies have explored how biodiversity influences transmission, particularly the *dilution effect*, where greater vertebrate species richness reduces pathogen prevalence by diverting tick blood meals to non-competent hosts [15, 62]. For instance, LoGiudice *et al*. [18] developed a mechanistic host-community model to show that biodiversity lowers the density of infected *I. scapularis* nymphs. Furthermore, Lou *et al*. [45] used a mechanistic model to show that seasonal variations and biodiversity reduce infection risk by distributing the pathogen among multiple host species. Spatial models have also been developed and used to examine how landscape structure influences Lyme disease spread. In particular, Caraco *et al*. [64] used a reaction-diffusion model to show that tick invasion rates and human exposure peak under intermediate tick mortality and host availability. Zhang *et al*. [49] incorporated co-feeding transmission to highlight its role in disease persistence, while Vindenes *et al*. [50] used a seasonal matrix population model to explore how tick life histories shape population dynamics. Shaw *et al*. [63] examined how habitat fragmentation alters tick-host interactions, showing that landscape connectivity and green space availability strongly influence pathogen prevalence. Network and agent-based models have also been used to study various aspects of the tick and disease dynamics, such as host movement, human exposure risk, and assessment of intervention strategies. For instance, Savage *et al*. [61] used a contact network model to show that seasonal fluctuations in human host availability impact nymphal infection prevalence. Nguyen *et al*. [51] also used a network metapopulation model to show that human mobility between endemic and non-endemic regions amplifies localized outbreaks of Lyme disease. Foster *et al*. [60] developed an agent-based model that integrates climate variability and host dispersal, and showed that targeted interventions, such as tick suppression and deer population management, effectively reduce disease incidence. Most of the aforementioned modeling studies have focused on the population dynamics of *I. scapularis* and Lyme disease in mice. In other words, these modeling studies did not focus on Lyme disease in humans. This forms one of the key aims of the current study, as described below.

The main objective of the current study is to investigate the influence of climate variability (as measured in terms of changes in local temperature) on the geospatial dynamics of *I. scapularis* and Lyme disease transmission in the U.S. state of Maryland. The objective will be achieved by the development, calibration, analysis, and simulation of a novel mechanistic model for the tick-host-pathogen dynamics associated with Lyme disease transmission and control in Maryland. The model will be calibrated and validated using both historical Lyme disease case data and fine-scale temperature data. The model, which explicitly accounts for the temperature-dependent tick-host interactions and tick dynamics, will be used to assess the impact of temperature fluctuation on the geospatial distribution of ticks and Lyme disease in the state of Maryland, as well as to assess the population-level impact of control and mitigation strategies against the disease. The paper is organized as follows. The model is formulated in Section 2. Its basic qualitative properties, as well as asymptotic stability of its equilibria, are also presented in this section. The model is calibrated with observed data in Section 2.4. Global sensitivity analysis is carried out in Section 2.8 to determine the parameters of the model that have the most influence on the disease dynamics. Numerical simulations are reported in Section 3, and concluding remarks and discussion are presented in Section 4.

## 2 Formulation of Lyme disease transmission model

The proposed model to be developed in this section monitors the transmission dynamics of Lyme disease within the tick, mice and human population in the U.S. state of Maryland. The model will incorporate key biological features associated with the tick-host-pathogen interactions, such as blood meal feeding, immature dynamics (eggs-larvae-nymphs-adult), and the effect of temperature on the tick-host-pathogen dynamics. The model is formulated as below.

### 2.1 State variables of the model

The total population of *I. scapularis* ticks at time *t*, denoted by *N*_*T*_ (*t*), is subdivided into the total subpopulations of larvae (denoted by *N*_*L*_(*t*)), nymphs (denoted by *N*_*N*_ (*t*)) and adult (*N*_*A*_(*t*)). Further, the total population of nymphs is subdivided into susceptible (*S*_*N*_ (*t*)) and infected (*I*_*N*_ (*t*)) at time *t*, so that *N*_*N*_ (*t*) = *S*_*N*_ (*t*) + *I*_*N*_ (*t*). Hence, the total ticks population at time *t* is given by

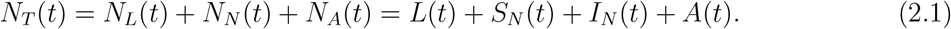

Similarly, the total population of mice at time *t*, denoted by *N*_*M*_ (*t*), is subdivided into the subpopulations of susceptible mice (*S*_*M*_ (*t*)) and infected mice (*I*_*M*_ (*t*)), so that

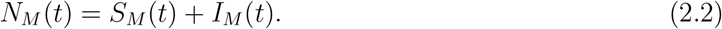

Finally, the total human population at time *t*, denoted by *N*_*H*_(*t*), is stratified into the subpopulations of susceptible (*S*_*H*_ (*t*)), exposed (*E*_*H*_(*t*)), infectious (*I*_*H*_(*t*)), hospitalized (*H*_*H*_(*t*)), and recovered (*R*_*H*_(*t*)) humans, so

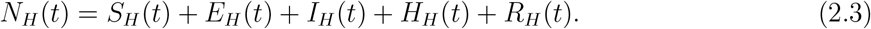

#### 2.1.1 Equations for the dynamics of ticks

Adult ticks primarily feed from deer to lay eggs, which later mature to become larvae. Larvae are generated at a rate 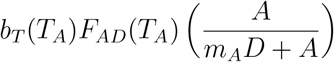,where *b*_*T*_ (*T*_*A*_) is the temperature-dependent number of eggs that hatch to larvae, *F*_*AD*_(*T*_*A*_) is the feeding rate of adult ticks on deer, *m*_*A*_ is the half *m* saturation constant for deer abundance, 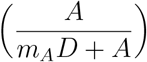 is the Holling Type II functional response accounting for the fact that adult feeding rate is dependent on the availability/abundance of deer, and *T*_*A*_(*t*) represents the local air/ambient temperature at time *t* (which is assumed to be continuous, bounded, positive and periodic [67, 68]). Larvae feed on mice (susceptible or infected). Larvae mature to susceptible or infected nymphs after successfully taking a blood meal from a susceptible or infected mouse, respectively. Let 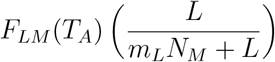 be the rate at which larvae feed from mice, where *F*_*LM*_ (*T*_*A*_) is the temperature-dependent feeding rate of larvae on mice, the term in parentheses is the Holling Type II functional response accounting for the fact that the feeding success depends on the density of the host (mice), with *m*_*L*_ representing the half saturation constant for availability of mice to the larvae. Larvae that successfully feed on susceptible mice (*S*_*M*_) mature to become susceptible nymphs (at the rate 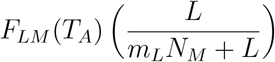).Those that fed on infected mice (*I* _*M*_) mature to become infected nymphs at a rate 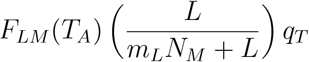 (where 0 *< q* − 1 is the probability of disease transmission to larvae by infected mice; thus, larvae that feed on infected mice fail to acquire the infection and become susceptible nymph, with probability 1− *q*_*T*_). Larvae that feed on infected mice are known to have very high likelihood of acquiring the infection (i.e., *q*_*T*_ ≈ 1 [69]). Larvae, nymphs, and adult ticks are lost due to natural causes such as predation (being eaten by predators such as birds and rodents), environmental stress (extreme temperatures, humidity changes, or habi-tat disturbances), resource depletion (lack of food or hosts for feeding), and physiological aging (the natural decline in bodily functions over time) at a rate *µ*_*L*_, *µ*_*N*_, and *µ*_*A*_, respectively [70]. Susceptible nymphs (*S*_*N*_) mature to become adult ticks (after successfully taking a blood meal from mice at a rate 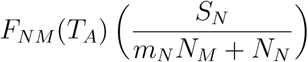,where *F*_*NM*_ (*T*_*A*_) be the rate at which susceptible nymphs feed on mice, where *F*_*NM*_ (*T*_*A*_) is the temperature-dependent feeding rate of nymphs on mice, and the term in parentheses represents the Holling Type II functional response accounting for host (mice) availability, and *m*_*N*_ is the half saturation constant for availability of mice to the nymphs. Similarly, susceptible nymphs feed on humans, and mature to become adult ticks, at a rate 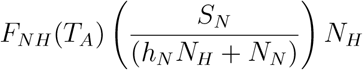, where *F*_*NH*_(*T*_*A*_) is the temperature-dependent nymph feeding rate on humans, and *h*_*N*_ is the half saturation constant for availability of humans to the nymphs. Infected nymphs (*I*_*N*_) feed on mice and mature to become adult ticks at a rate. The population of infected nymphs (*I*_*N*_) is generated by the feeding of larvae on infected mice at the rate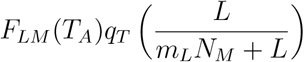. Similar to suscep-tible nymphs, infected nymphs also feed on both mice and humans to become adult ticks (at the rates 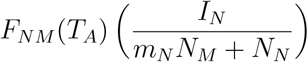 and 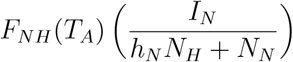,respectively). Adult ticks (*A*) emerge when both susceptible and infected nymphs successfully feed (on both mice and humans) and molt into the adult stage. Susceptible nymphs feeding on mice and humans contribute to the production of adult ticks at the rates 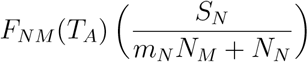 and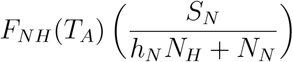, respectively. Simi-larly, infected nymphs contribute to the adult tick population at the rates 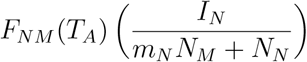 and 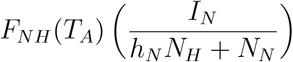, respectively. Adult tick feed on deer (to acquire the blood meal needed for the development of eggs) at a rate 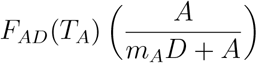, where *F*_*AD*_ (*T*_*A*_) is the feeding rate of adult ticks on deer, and the term in parentheses accounts for the saturation effect due to deer abundance. Based on the above derivations and assumptions, the equations for the dynamics of ticks and Lyme disease are given by the following deterministic system of nonlinear differential equations:

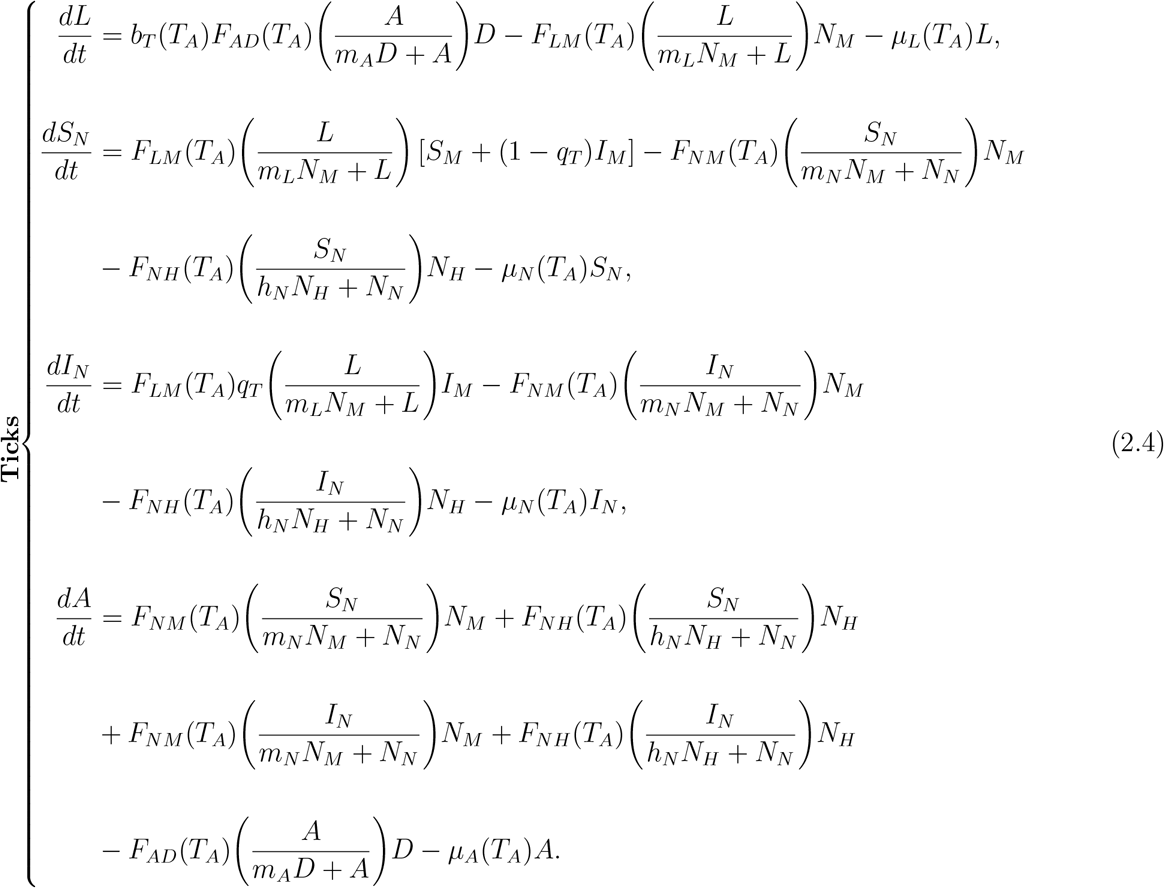

### 2.1.2 Equations for the dynamics of mice

Mice serve as key reservoir hosts in the Lyme disease transmission cycle, acquiring infection from the bite of infected nymphs, and subsequently infecting larvae that feed on them (the infected larvae do not transmit infection to the mice; they only molt to become infected nymphs [12]. Mice offsprings are produced at a temperature-dependent rate *b*_*M*_ (*T*_*A*_), which accounts for litter size and is assumed to be proportional to mouse population density *N*_*M*_ (*t*) [47]. Susceptible mice acquire Lyme disease infection following an effective bite by an infected nymph (*I*_*N*_) at a nonlinear rate 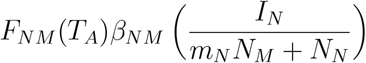, where *FNM* (*T*_*A*_) is the temperature-dependent feeding rate of nymphs on mice, *β*_*NM*_ is the rate of infection of susceptible mice *per* bite from an infected nymph and 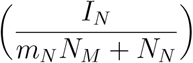 is the Holling Type II functional response accounting for the saturation in feeding behavior of nymphs on mice) [18, 71]. Mice (susceptible and infected) are lost naturally at a temperature-dependent rate *µ*_*M*_ (*T*_*A*_), and *via* additional density-dependent effects (accounting for intra-species competition for resources), at a rate *δ*_*M*_ *N*_*M*_ */K*_*N*_ (*T*_*A*_), where *K*_*M*_ (*T*_*A*_) is the temperature-dependent carrying capacity for mice, accounting for ecological pressures such as food availability, predation, and habitat conditions for mice [72, 73]. Infected mice do not recover from *B. burgdorferi* infection, but serve as pathogen reservoirs, thereby sustaining the Lyme disease enzootic transmission cycle. [18]. It follows, based on the above derivations and assumptions, that the equations for Lyme disease transmission in mice are given by:

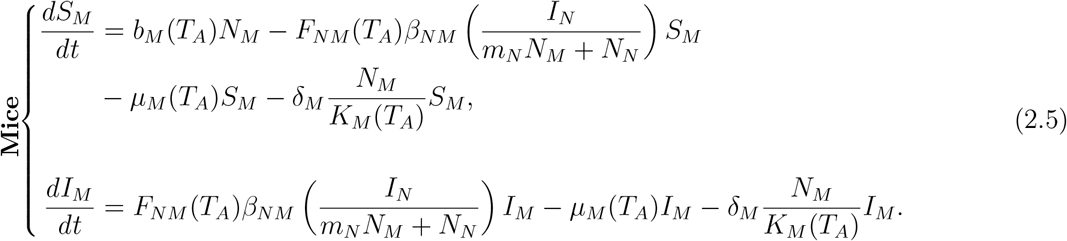

### 2.1.3 Equations for the dynamics of humans

Humans are incidental hosts in the Lyme disease transmission cycle and do not contribute to the enzootic maintenance of *B. burgdorferi* [12]. Specifically, humans exclusively acquire Lyme disease infection through bites from infected nymphs, and they do not contribute to further transmission of the disease. The population of humans in the community is increased by recruitment (birth or immigration) at a rate (assumed constant) Π_*H*_ and by the loss of infection-acquired immunity of recovered individuals, at a rate *ψ*_*H*_. Susceptible humans (*S*_*H*_) acquire Lyme disease infection following an effective bite from an infected nymphs (*I*_*N*_), at a rate 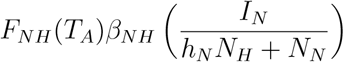 where *F*_*NH*_(*T*_*A*_) is the temperature-dependent feeding rate of nymphs on humans and *β*_*NH*_ is the probability of infection *per* bite from an infected nymph to a susceptible human. Humans in all epidemiological compartments die naturally at a rate *µ*_*H*_. Exposed individuals (in the *E*_*H*_ class) developed clinical symptoms of Lyme disease at a rate *σ*_*H*_, and symptomatic individuals transition out of the *I*_*H*_ class at a rate *τ*_*H*_ (where a proportion, 0 *< g*_*H*_ *<* 1, recovers, and the remaining proportion, 1 − *g*_*H*_, are hospitalized). Hospitalized individuals recover at a rate *γ*_*H*_. Since death due to Lyme disease in the United States is very rare (only 9 deaths were recorded over the last 30 years [2]), it is assumed that humans do not suffer Lyme disease-induced mortality. Based on the above derivations and assumptions, the equations for the dynamics of Lyme disease in humans are:

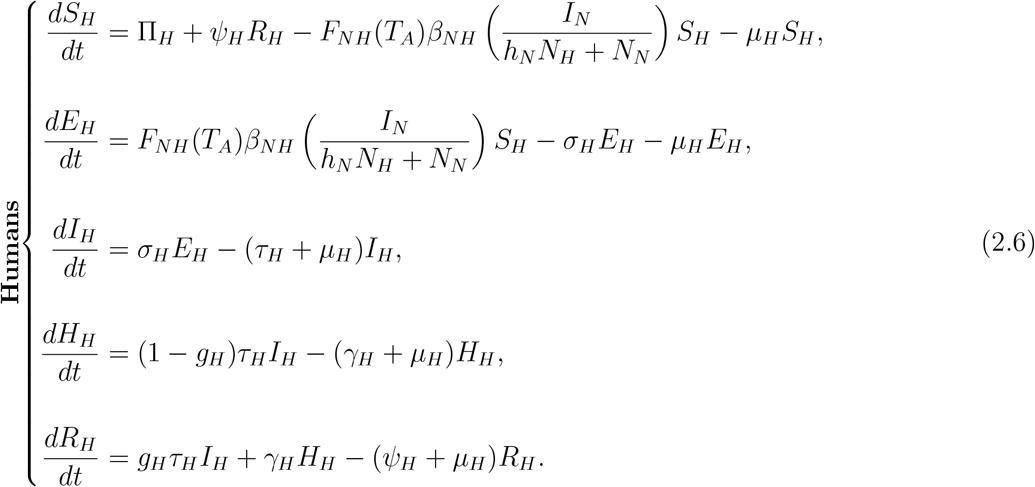

The transmission dynamics of Lyme disease among ticks, mice, and humans are illustrated in the model flow diagram shown in Figure 2, corresponding to the system of equations (2.4)–(2.6). Table 21 summarizes the state variables and their descriptions. The parameters of the model (2.4)– (2.6) are described in Table 22; the functional forms of the temperature-dependent parameters are derived in Section 2.2.

**Table 21:** Description of state variables of the model {(2.4)-(2.6)}.

| State variable | Description |
| --- | --- |
| $L$ | Density of larvae |
| $S_N$ | Density of susceptible nymphs |
| $I_N$ | Density of infected nymphs |
| $A$ | Population of adult ticks tick |
| $S_M$ | Population of susceptible mice |
| $I_M$ | Population of infected mice |
| $S_H$ | Population of susceptible humans |
| $E_H$ | Population of exposed humans |
| $I_H$ | Population of infected humans |
| $H_H$ | Population of hospitalized humans |
| $R_H$ | Population of recovered humans |
| $D$ | Population density of deer |

**Figure 2:**
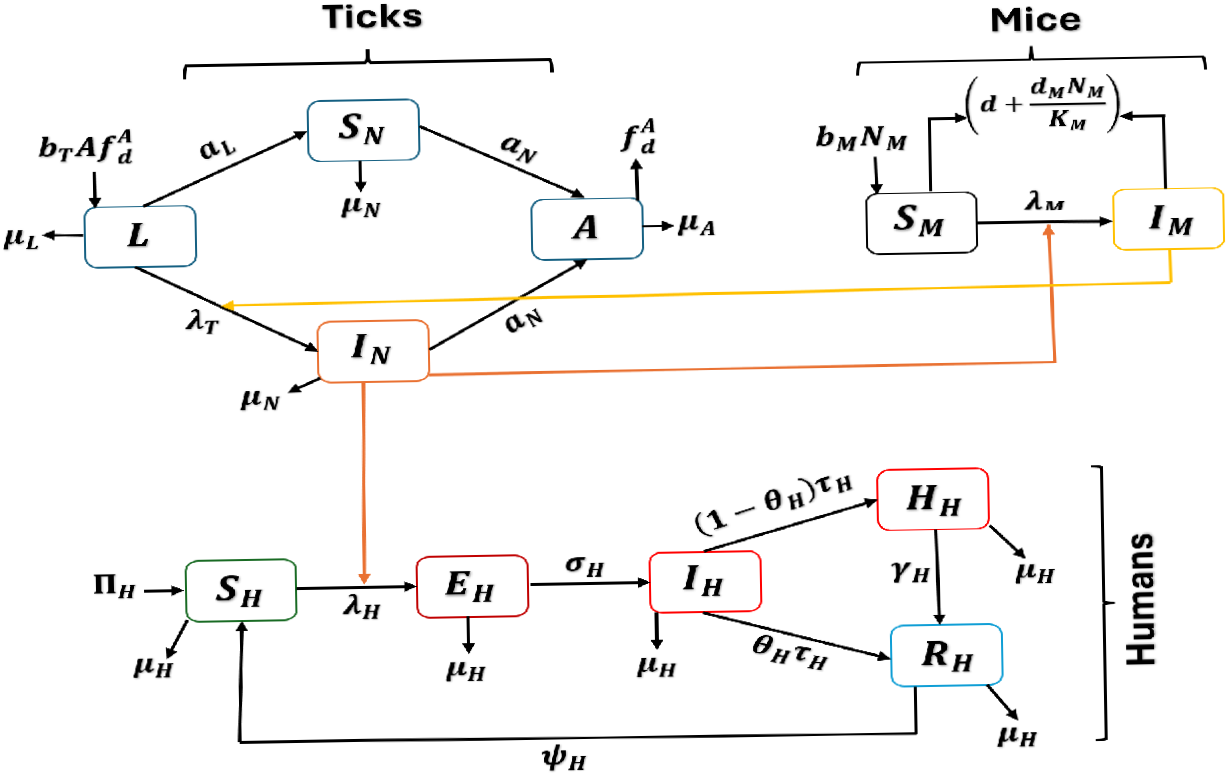
Flow diagram of the Lyme disease transmission model {(2.4)-(2.6)}, showing the dy-namics between ticks, mice and humans. **Notations:** 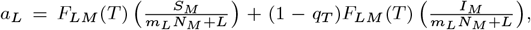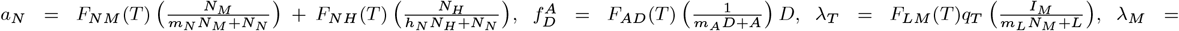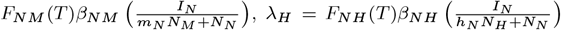 The temperature-dependent feeding rate param-eters of the model are described in detail in Table **??** of Supplementary Material **??**.

Some of the main assumptions made in the formulation of the model {(2.4)-(2.6)} are:

i. Homogeneous mixing: it is assumed that the populations of ticks, mice, humans, and deer are well-mixed (so that the host species have equal likelihood of mixing among themselves and with other species).
ii. No spatial heterogeneity for ticks habitats, landscape, and Lyme disease exposure risk: although heterogeneity in ticks population abundance and transmission dynamics exists (between urban and forested areas, for example [74, 75]), such spatial heterogeneity is not explicitly accounted for in the model for mathematical tractability.
iii. No vertical transmission of *B. burgdorferi* from infected adult female ticks to their offspring [76] (all ticks larvae molt into nymphs, regardless of the infection status of the adult ticks that lay eggs that hatch into the larvae [12]). There is also no vertical transmission of *B. burgdorferi* from infected mother to her child [77].
iv. No transmission of *B. burgdorferi* to susceptible ticks during co-feeding with infected ticks on a competent host. Although an infected tick co-feeding with susceptible ticks on a competent host (mice) can transmit *B. burgdorferi* to the host, who, in turn, can transmit *B. burgdorferi* to the co-feeding susceptible, this process is not accounted for in the model because it is rare (e.g., the two ticks need to be within 1 to 2 cm radius of each other on the host [78–80] and that the susceptible tick has to be on the host for an extended period of time, related to the incubation or latency period of the host), compared to the direct (systemic) infection of the susceptible tick that is either feeding alone on the host, or co-feeding with others at a distant location (longer than 2 cm) on the host [44, 49, 81, 82].
v. Humans are dead-end hosts: Humans acquire Lyme disease infection through tick bites but do not transmit *B. burgdorferi* to ticks or other hosts [82].
vi. No direct tick-to-tick transmission: Ticks acquire infection only through feeding on infectious hosts and do not transmit *B. burgdorferi* directly to each other [47].
vii. Mice are carriers, and do not die of Lyme disease. Further, it is assumed that humans do not succumb to Lyme disease infection.
viii. Adult ticks are not stratified according to infection status (susceptible or infected). This is because of the fact that adult ticks primarily feeds on deer, and deer do not acquire Lyme disease infection [47]. It is assumed that the deer population is always present in the community [83], so that adult ticks can always find a blood meal.
ix. We assume that *B. burgdorferi* establishes rapidly in both ticks and mice following infection, allowing us to omit an exposed compartment in both ticks and mice populations, as supported by experimental evidence showing systemic infection in mice within 2–4 days and prompt midgut colonization in ticks [84, 85].
x. Infectious nymphs remain infectious for life, transmitting *B. burgdorferi* throughout their stage, emerging in late spring (May–July) and maturing into adults by fall (September–November). Eggs are laid in spring (April–June), hatching into larvae in summer (July–September), which molt into nymphs the following spring. Adults play a minimal role in *B. burgdorferi* amplification [65, 82].

**Table 22:** Description of the parameters of the model {(2.4)-(2.6)}.

| Parameter | Description |
| --- | --- |
| **Ticks** |  |
| $q_T$ | Probability of Lyme disease transmission to larvae by infected mice |
| $b_T(T_A)$ | Hatching rate of eggs |
| $F_{LM}(T_A)$ | Feeding rate of larvae on mice |
| $\mu_L(T_A)$ | Natural mortality rate of larvae |
| $\mu_N(T_A)$ | Natural mortality rate of nymphs |
| $\mu_A(T_A)$ | Natural mortality rate of adult ticks |
| **Mice** |  |
| $b_M(T_A)$ | Birth rate of mice |
| $\beta_{NM}$ | Rate of infection of susceptible mice from an infected nymph |
| $F_{NM}(T_A)$ | Feeding rate of nymphs on mice |
| $m_L$ | Half saturation constant for availability of mice to larvae |
| $m_N$ | Half saturation constant for availability of mice to nymphs |
| $K(T_A)$ | Carrying capacity of mice |
| $\mu_M(T_A)$ | Natural mortality rate of mice |
| $\delta_M$ | Density-dependent mortality rate of mice |
| **Humans** |  |
| $\Pi_H$ | Recruitment (birth or immigration) rate of humans |
| $\mu_H$ | Natural death rate for humans |
| $\beta_{NH}$ | Rate of transmission from infected nymph to susceptible humans |
| $F_{NH}(T_A)$ | Feeding rate of nymphs on humans |
| $h_N$ | Half saturation constant for availability of humans to nymphs |
| $\sigma_H$ | Progression rate from exposed to infectious humans |
| $g_H$ | Proportion of hospitalized humans |
| $\tau_H$ | Transition rate out of the symptomatic class |
| $\gamma_H$ | Recovery rate of humans |
| $\psi_H$ | Rate of loss of infection-acquired immunity |
| **Deer** |  |
| $F_{AD}(T_A)$ | Feeding rate of adult ticks on deer |
| $m_A$ | Half-saturation constant for adult ticks feeding on deer |

The model {(2.4)-(2.6)} is an extension of several models for the ticks-host-pathogen dynamics associated with Lyme disease transmission, such as those presented in [23, 44–47, 49, 53, 60–62, 86], by, *inter alia*:

i. Explicitly incorporating human dynamics (with humans as dead-end hosts). This is not accounted for in several models for Lyme disease transmission dynamics, such as the models in [47, 49, 49, 53, 60, 61].
ii. Explicitly accounting for the effect of local temperature variations on the ticks-host-pathogen dynamics, particularly ticks feeding success, survival, development, questing activity, and the hosts’ population dynamics (this was not included in the models presented in [54]). In addition to adding realism to the ecological component of the model (since temperature affects all aspects of the lifecyle of *I. scapularis* and the *B. burgdorferi* bacteria), explicitly adding temperature effects enable the realistic qualitative assessment of the population abundance of ticks, its hosts, and Lyme disease in a population under various climate change projections.
iii. Explicitly using nonlinear feeding rates for ticks: The model uses Holling Type II functional responses to explicitly account for the monotone (and saturation) nature of feeding rates of ticks on the competent hosts (linear feeding rates were used in [46, 86]). Furthermore, although such functional responses were used for the feeding rates in the model developed in [47], the model in [47] did not include humans as a host for Lyme disease (unlike in our study).

### 2.2 Functional forms of the thermal response functions of the model

The model {(2.4)-(2.6)} contains several time-dependent parameters associated with the impact of temperature on the lifecycle of ticks and the ecology of the mice (such as the feeding, reproduction, and survival of the ticks species (*I. scapularis*), as well as reproduction, mortality, and carrying capacity of mice species (*P. leucopus*); these, collectively, shape *B. burgdorferi* transmission dynamics [23, 44, 46, 47]). For instance, it is known that ticks questing of blood meals peaks at 18°C, while extreme temperatures reduce tick survival and alter host population structure [44]. The functional forms of the thermal response functions of the model will now be formulated, based on data collected from empirical studies [87], as described below (the functional forms of the time-dependent parameters are depicted in Figure 3).

**Figure 3:**
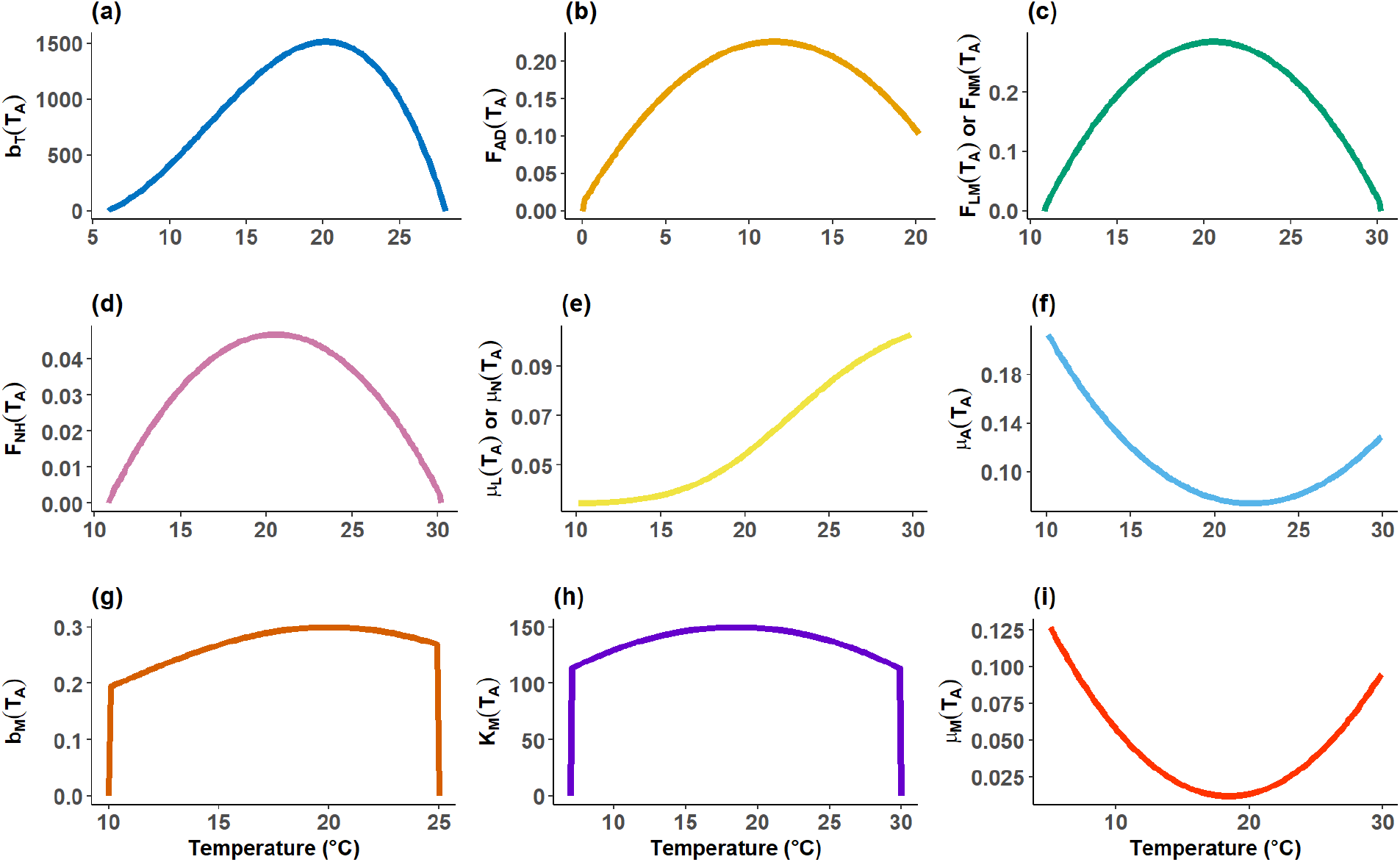
Profiles of the temperature-dependent parameters of the model {(2.4)–(2.6}) for the state of Maryland, generated by evaluating the functional forms in Section 2.2 with mean daily temperature during a typical tick season in Maryland. (**a**) Hatching rate of eggs (*b*_*T*_ (*T*_*A*_)); (**b**) feeding rate of adult ticks on deer (*F*_*AD*_(*T*_*A*_)); (**c**) feeding rates of larvae and nymphs on mice (*F*_*LM*_ (*T*_*A*_) and *F*_*NM*_ (*T*_*A*_), respectively, assumed to have the same functional form); (**d**) feeding rate of nymphs on humans (*F*_*NH*_(*T*_*A*_)); (**e**) natural mortality rates of larvae and nymphs (*µ*_*L*_(*T*_*A*_) and *µ*_*N*_ (*T*_*A*_), respectively; assumed to have the same functional form)(**f**) natural mortality rate of adult ticks (*µ*_*A*_(*T*_*A*_)); (**g**) birth rate of mice (*b*_*M*_ (*T*_*A*_)); (**h**) carrying capacity of mice (*K*_*M*_ (*T*_*A*_)); and (**i**) natural mortality rate of mice (*µ*_*M*_ (*T*_*A*_)). In generating these thermal response curves, all parameters in the functional forms of the temperature-dependent parameters that are not dependent on temperature are fixed at their baseline values given either in Table **??** of Supplementary Material **??** or in the main text under Section 2.2.

#### Hatching rate of eggs *b*_*T*_ (*T*_*A*_)

Ambient temperature greatly affects the reproductive success of *I. scapularis* (*vis a vis* the oviposition rate and survival probability of eggs). For example, while warmer temperatures (within an optimal range) promote the successful hatching of eggs, extremely high temperatures increase the desiccation risk of eggs. Furthermore, extremely cold temperatures impair embryonic development [87, 88]. The hatching rate of eggs (to become larvae) is defined as:

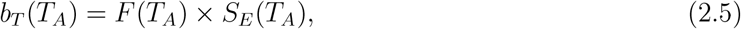

where *F* (*T*_*A*_) represents the temperature-dependent fecundity rate of mated adult female ticks (which is the product of the oviposition rate of the adult female tick and the number of eggs laid *per* oviposition) and *S*_*E*_(*T*_*A*_) denotes the survival probability of the eggs laid by the tick. Following [87, 88], the fecundity function takes the form (which peaks at around *T*_*A*_ = 17°C, in line with empirical studies conducted in [42, 87]):

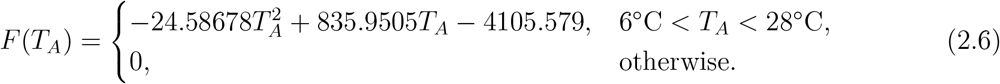

Similarly, the egg survival probability is given by:

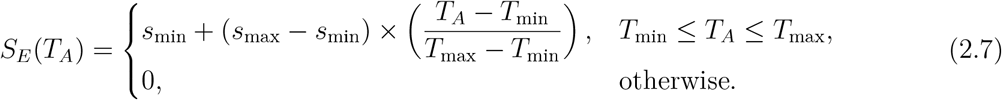

where *T*_min_ = 6°C, *T*_max_ = 28°C, and *s*_min_ = 0.1, *s*_max_ = 0.8 [44, 46].

#### Tick feeding rates (*F*_*i,j*_(*T*_*A*_))

Tick feeding behavior is regulated by temperature, which influences questing activity, metabolic rates, and host-seeking success. For instance, while increased temperature (within a certain suitable range) enhances feeding efficiency, tick mobility declines at extreme (high or low) temperatures due to desiccation stress (such as insufficient nutrients for ticks and availability of hosts) [23, 44]. Following [44, 46], the temperature-dependent feeding rate for a tick at stage *i* (where *i* = larval, nymphal, or adult stage) feeding on host *j* (where *j* = mouse, human, or deer) is given by:

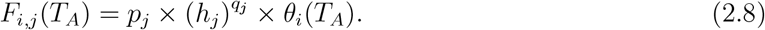

The parameter 0 *< p*_*j*_ ≤ 1 represents the intrinsic host-questing efficiency of ticks (larvae, nymphs and adult), and 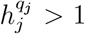 is a measure of the host population density (which may correspond to the total populations of mice (*N*_*M*_), humans (*N*_*H*_), or deer (*D*) in the environment or community), with 0 *< q*_*j*_ ≤ 1 serving as a scaling factor that adjusts for the effect of the availability (or lack thereof) of host type *j* on the feeding success of ticks at stage *i*. Additionally, *θ*_*i*_(*T*_*A*_) is the temperature-dependent questing rate for ticks at stage *i*. Specifically, the host-questing of larvae (*θ*_*L*_(*T*_*A*_), obtained by fitting the experimental data collected in [87], is given by (this formulation was also used in [42, 46, 48, 87, 88])

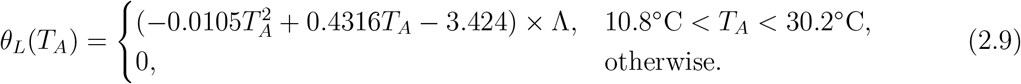

where,

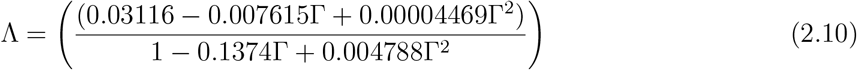

and Γ accounts for seasonal variation in tick activity influenced by photoperiod (Γ), estimated with an annual mean of approximately 12 hours across North America [89, 90]. Although the precise impact of photoperiod on *I. scapularis* activity remains unsettled [43], empirical data indicate peak questing occurs at temperatures between 20–25°C [42, 46]. It is worth mentioning that in this study, we make the simplifying assumption that the parameters for the temperature-dependent feeding rates for larvae on mice (*F*_*LM*_ (*T*_*A*_)) and nymphs on humans (*F*_*NH*_ (*T*_*A*_)) are the same (i.e., they share the same values of the parameters 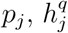,and *θ*_*i*_; so that= *F*_*LM*_ (*T*_*A*_) = *F*_*NH*_(*T*_*A*_)). This is owing to their comparable host-seeking behavior and metabolic constraints during their developmental stages, as supported by prior empirical studies [42, 87]. Following [88], the host-questing for nymphs (*θ*_*N*_ (*T*_*A*_)) is assumed to be the same as that of larvae (i.e., *θ*_*N*_ (*T*_*A*_) = *θ*_*L*_(*T*_*A*_)). Finally, following [42, 88], the host-questing rate for adult ticks (*θ*_*A*_(*T*_*A*_)) is given by:

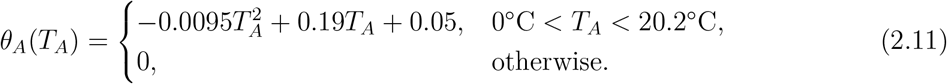

#### Natural mortality rates for ticks (*µ*_*L*_(*T*_*A*_), *µ*_*A*_(*T*_*A*_))

Tick mortality follows a temperature-dependent pattern, with optimal survival at intermediate temperatures and increased mortality under extreme heat or cold due to desiccation or reduced overwintering survival [43, 88, 90]. In the absence of empirical data for the effect of temperature on the natural mortality for ticks, we adopt the functional forms used for the effect of temperature on other disease vectors (such as mosquitoes) to model these rates. In particular, following the formulations in [67, 91] (for malaria mosquitoes), the temperature-dependent larval and nymphal mortality rate is given by (larvae and nymphs exhibit similar mortality responses to temperature, given their shared ecological constraints. Hence, these rates are assumed to be the same for the two tick stages). That is,

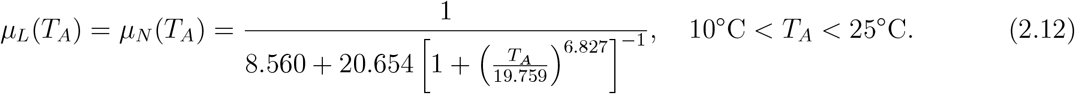

This function captures the non-linear mortality rate for larvae and nymphs, with peak survival at intermediate temperatures [67, 91].

Adult ticks are more resilient due to their thicker cuticle and reduced surface-area-to-volume ratio, but remain susceptible to desiccation and metabolic stress [92, 93]. Following [23, 67, 91], the temperature-dependent natural mortality rate for adult ticks is given by (this function shows a profile that is consistent with observed survival patterns for ticks as temperature increases):

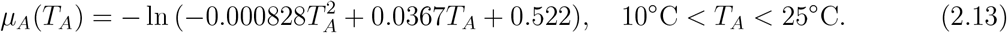

#### Birth rate of mice (*b*_*M*_ (*T*_*A*_))

Temperature affects various aspects of the mice reservoir host (*P. leucopus*), such as reproduction, carrying capacity, and survival [90, 94]. For instance, it is known that the birth rate of mice peaks at a certain temperature range that supports resource abundance and physiological stability of the mice (notably surviving anti-ticks control measures that affect mice reproduction, such as acaricide-treated bait stations, thermacell tick control tubes, rodenticide, and permethrin-treated bait stations [95–97]). The temperature-dependent birth rate of mice is given by:

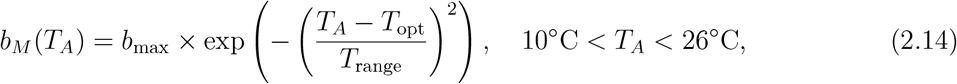

where *b*_max_ = 0.3 *per* day, *T*_opt_ = 20°C, and *T*_range_ = 15°C [94, 98, 99].

#### Carrying capacity of mice (*K*_*M*_ (*T*_*A*_))

Temperature affects the population density (or environmental carrying capacity) of mice due to its impact on resource constraints (such as food availability, habitat suitability, and reproduction) [97, 99]. Warmer temperatures can increase access to food sources—such as beetles and caterpillars, which are part of the mouse diet—and improve habitat conditions, but may also accelerate spoilage. Higher temperatures tend to extend breeding seasons and shorten birth intervals, although extreme heat can reduce reproductive success [99]. Temperature also influences predation and mortality, further shaping population dynamics.

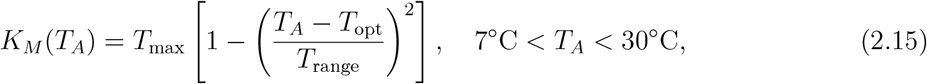

where *T*_max_ is the maximum temperature for mice survival, *T*_opt_ is the optimal temperature for mice survival and *T*_range_ is the average of the minimum and maximum daily temperature in the environment [99].

#### Natural mortality rate of mice (*µ*_*M*_ (*T*_*A*_))

Temperature affects the mortality of mice, where the rate is maximized at extreme temperatures. The temperature-dependent natural mortality rate for mice is given by:

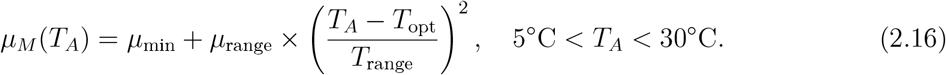

where *µ*_min_ = 0.012 *per* day, *µ*_range_ = 0.4 *per* day [47, 98].

In order to estimate when the key ecological processes associated with peak seasonal variations of ticks and their reservoir host in Maryland (i.e., when they reach peak abundance), the functional forms of the 11 temperature-dependent parameters of the model {(2.4)–(2.6)}, given by equations {(2.5)–(2.16)}, are evaluated using the mean daily temperature data for the state of Maryland for a given year. Figure 4 depicts the monthly profiles of 9 of the 11 of these temperature-dependent(since we set *F*_*NM*_ (*T*_*A*_) = *F*_*LM*_ (*T*_*A*_) and *µ*_*N*_ (*T*_*A*_) = *µ*_*L*_(*T*_*A*_) above, in line with the empirical studies in [42, 87, 88]). This figure shows that, for a given year in Maryland, the key ecological processes for the tick-reservoir dynamics tend to generally peak during the spring and summer months. Specifically, both the egg hatching rate (*b*_*T*_ (*T*_*A*_)) and birth rate of mice (*b*_*M*_ (*T*_*A*_))—panels **(a)** and **(b)**—peak between May and June, followed by a decline beginning in July and a secondary, smaller peak in September. However, there is a decline until the end of the year, and the rates remain relatively low through the new year. These profiles for *b*_*T*_ (*T*_*A*_) and *b*_*M*_ (*T*_*A*_) suggest the presence of seasonal synchrony between larval tick emergence and juvenile mouse mice availability. The feeding rates—*F*_*AD*_(*T*_*A*_) (for adult ticks on deer), *F*_*LM*_ (*T*_*A*_) (for larvae on mice), *F*_*NM*_ (*T*_*A*_) (for nymphs on mice), and *F*_*NH*_(*T*_*A*_) (for nymphs on humans)—shown in panels **(b)**–**(d)**—also exhibit strong seasonal trends. For instance, *F*_*AD*_(*T*_*A*_) and *F*_*LM*_ (*T*_*A*_) (panels **(b)** and **(c)** attain their maximum during June–August, while *F*_*NH*_(*T*_*A*_) (panel **(d)**) display bimodal patterns that closely mirror those of *b*_*T*_ (*T*_*A*_) and *b*_*M*_ (*T*_*A*_). This resemblance likely reflects ecological coupling between host availability and tick activity—particularly during periods of high vertebrate host abundance and reproductive activity [23, 33]. These patterns align with established host-seeking behaviors of the tick life stages and reveal the timing of increased contacts or interactions between ticks and their reservoir and accidental hosts.

**Figure 4:**
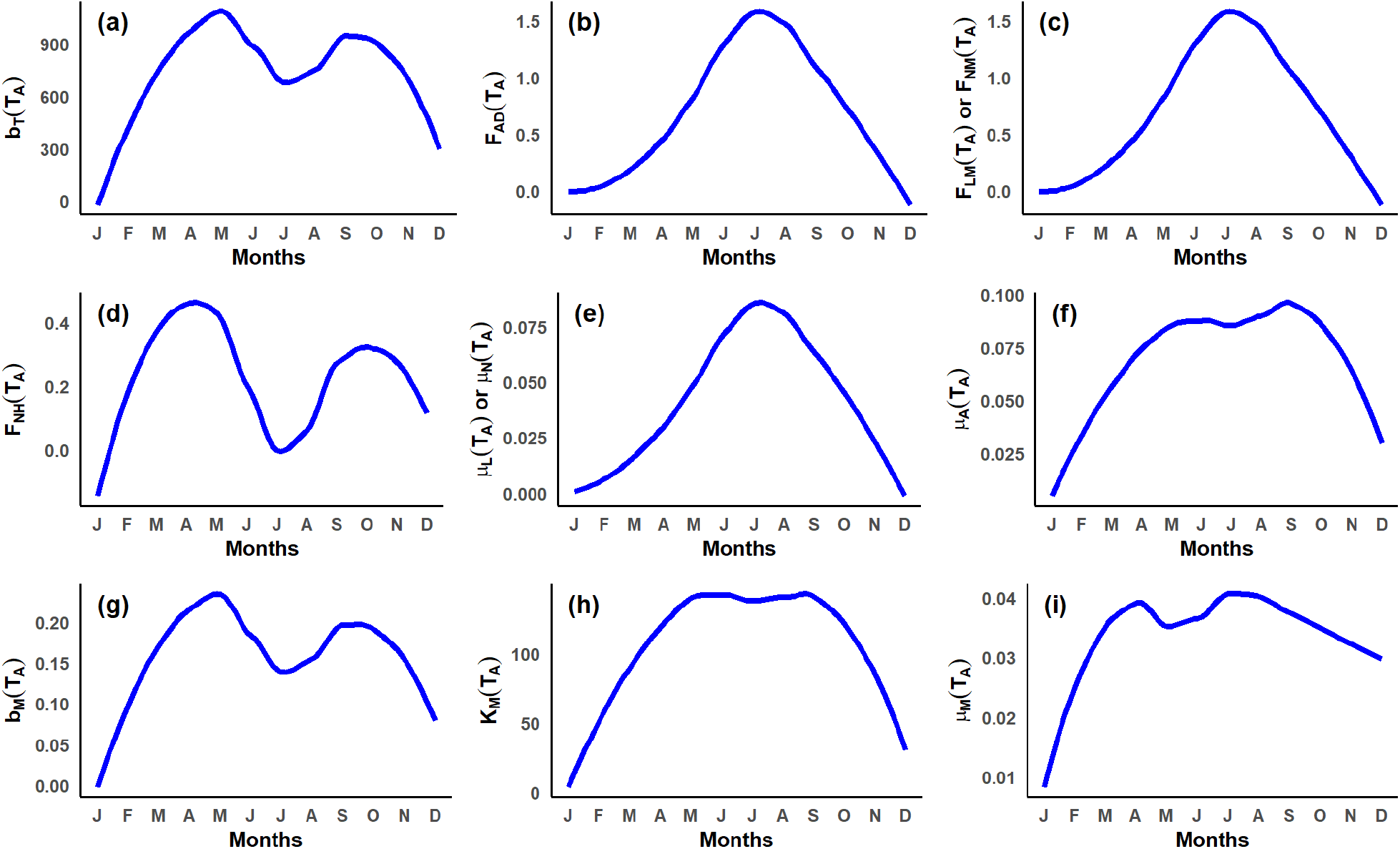
Seasonal profiles of the 11 temperature-dependent parameters of the model {(2.4)–(2.6)} generated using the functional forms in Section 2.2 and observed mean daily temperature for the state of Maryland during the year 2022 (available from [41, 102, 103]). Each panel gives the monthly profile of each of the temperature-dependent parameters (i.e., monthly averages of these parameters are plotted, based on averaging their daily values for the month). (**a**) Hatching rate of eggs (*b*_*T*_ (*T*_*A*_)); (**b**) feeding rate of adult ticks on deer (*F*_*AD*_(*T*_*A*_)); (**c**) feeding rates of larvae and nymphs on mice (*F*_*LM*_ (*T*_*A*_) and *F*_*NM*_ (*T*_*A*_), respectively, assumed to have the same functional form); (**d**) feeding rate of nymphs on humans (*F*_*NH*_(*T*_*A*_)); (**e**) natural mortality rates of larvae and nymphs (*µ*_*L*_(*T*_*A*_) and *µ*_*N*_ (*T*_*A*_), respectively; assumed to have the same functional form)(**f**) natural mortality rate of adult ticks (*µ*_*A*_(*T*_*A*_)); (**g**) birth rate of mice (*b*_*M*_ (*T*_*A*_)); (**h**) carrying capacity of mice (*K*_*M*_ (*T*_*A*_)); and (**i**) natural mortality rate of mice (*µ*_*M*_ (*T*_*A*_)). In generating these thermal response curves, all parameters in the functional forms of the temperature-dependent parameters that are not dependent on temperature are fixed at their baseline values given either in Table **??** of Supplementary Material **??** or in the main text under Section 2.2.

Figure 4 further shows that mortality rates for juvenile ticks—*µ*_*L*_(*T*_*A*_) and *µ*_*N*_ (*T*_*A*_)—are highest during July–August (panel **(e)**), while the mortality rate for adult ticks (*µ*_*A*_(*T*_*A*_)), depicted in panel **(f)**, has two peaks, one in April and another in September (with the latter peak slightly higher), suggesting increased mortality from heat-related stresses. These mortality rates remain relatively low from November through March until the end of the year, likely due to cooler ambient temperatures reducing metabolic demand and desiccation risk. However, the mortality rate for mice (*µ*_*M*_ (*T*_*A*_))—panel **(i)**—exhibits two distinct peaks: one in March–April and another in July–August. The early spring rise may reflect lingering effects of winter-related stress and limited resource availability, while the midsummer peak likely results from heat exposure, elevated predation risk, and intra-species competition during peak breeding activity. Mortality is lowest during January–February and November–December, potentially due to factors such as metabolic adaptations to cold. These patterns are consistent with documented seasonal vulnerabilities in *Peromyscus* species and other small mammals in temperate North America [100, 101]. The carrying capacity of mice (*K*_*M*_ (*T*_*A*_))—panel **(h)**—begins increasing gradually from January through March, with sharper growth in April, exhibiting two peaks, one in May and another in September, and declines markedly from October through December, particularly under extreme temperature conditions.

These seasonal profiles highlight critical ecological windows of elevated tick–host interactions that heighten Lyme disease risk in Maryland. In particular, the results depicted in Figure 4 show that tick activity and host abundance are maximized in Maryland during late spring and early summer, providing the optimal time period to intensify control efforts against the vector and the reservoir hosts (e.g., targeted rodent treatment, habitat modification, and public awareness campaigns on personal protection). In other words, this study identifies the optimal time window, during the annual cycle, within which public health control measures targeting ticks and their hosts should be intensified. Moreover, lower-effort control strategies may suffice during periods of naturally reduced tick and host activity, such as late summer and winter. Thus, these findings provide a data-driven scientific basis for timing of control efforts to maximize their impact on reducing the population abundance of ticks and reservoir hosts, as well as to maximize their impact in significantly reducing the geospatial spread and burden of Lyme disease in Maryland.

### 2.3 Existence and asymptotic stability of disease-free equilibria

In this section, conditions for the existence and asymptotic stability of the disease-free equilibria of the *autonomous* version of the model {(2.4)–(2.6)}, where all temperature-dependent parameters are evaluated at the fixed temperature *T*_*A*_ = 18°C (corresponding to the mean of monthly average temperatures during the tick season, usually April–October [41, 104, 105]), will be explored. The objective is to determine conditions, in parameter space, for the persistence or extinction of the *I. scapularis* ticks and *P. leucopus* mice populations in a local human environment. It is, first of all, convenient to define the following quantities:

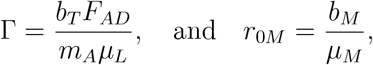

where Γ is the *tick larvae production number*, representing the average number of tick larvae produced by a single adult female tick that successfully feeds on deer, and *r*_0*M*_ is the *mice production number*, which measures the average number of mice offspring produced by an adult female mouse during its lifetime. The model {(2.4)–(2.6)} has a trivial disease-free equilibrium (TDFE), where no mice and ticks are present in the community, given by

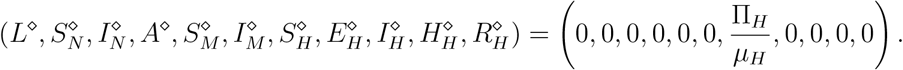

The result below can easily be established by linearizing the model {(2.4)–(2.6)} around the TDFE (as detailed in Supplementary Material **??**):

#### Theorem 2.1.

*The trivial disease-free equilibrium (TDFE) of the model* {(2.4)*–*(2.6)} *is locally-asymptotically stable whenever r*_0*M*_ *<* 1, *and unstable whenever r*_0*M*_ *>* 1.

The TDFE represents the ecological landscape where ticks and mice are not present in the environment (only humans are present), and no Lyme disease occurs in the human population. Although mathematically feasible (i.e., it always exists), this equilibrium is biologically-unrealistic in regions where Lyme disease is endemic, such as the Northeastern United States, where both the *I. scapularis* ticks and *P. leucopus* mice and other hosts (e.g., deer and humans) are present in abundance [41]. The model also has a nontrivial disease-free equilibrium with ticks and humans only present (i.e., no mice in the environment), given by

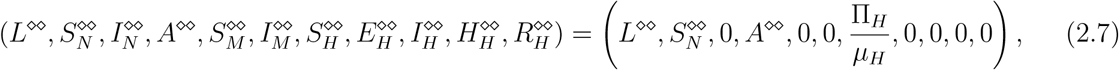

where 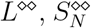, and *A*^−^ represent, respectively, the positive values of tick larvae, susceptible nymphs, and adult ticks at this equilibrium (it should be recalled that deer are assumed to always be present in the population, so that nymphs and adult ticks can always find a deer to take a blood meal from). Numerical simulations suggest that this equilibrium exists only when Γ *>* 1 (a condition associated with the persistence of ticks in the environment) and *r*_0*M*_ *<* 1 (ensuring the extinction of mice in the environment, in line with Theorem 2.1). The model also has a mice-humans-only nontrivial disease-free equilibrium (where ticks are not present in the environment), given by (this equilibrium exists only when *r*_0*M*_ *>* 1):

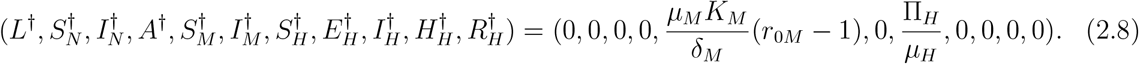

Finally, the model has a co-existence nontrivial disease-free equilibrium, where ticks, mice, and humans are present in the environment, given by (here, too, this equilibrium requires *r*_0*M*_ *>* 1, in addition to other conditions, for existence):

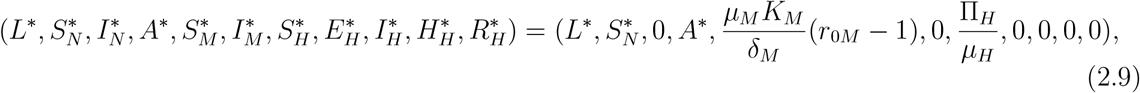

Where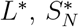, and *A*^*\**^ are the positive values of *L*(*t*), *S*_*N*_ (*t*) and *A*(*t*) at the non-trivial disease-free equilibrium (NDFE) and are obtained for each of the 24 Maryland counties by solving the following nonlinear system of equations (derived from the solutions of the model {(2.4)–(2.6)} at the NDFE):

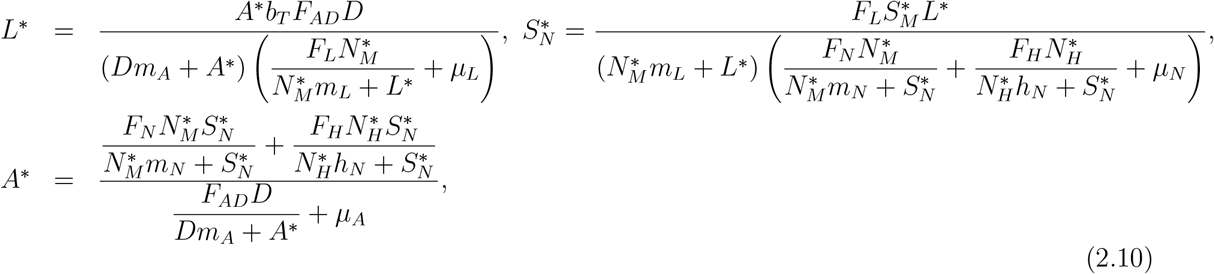

with 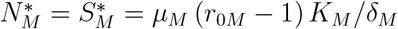,and 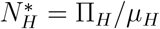. The local asymptotic stability of the co-existence disease-free equilibrium of the model {(2.4)–(2.6)}, with temperature fixed at *T*_*A*_(*t*) = 18°C, is explored using the next generation operator method [106, 107]. In particular, using the notation in [106], with the infected compartments of the model ordered as (*I*_*N*_, *I*_*M*_, *E*_*H*_, *I*_*H*_, *H*_*H*_), the non-negative matrix *F*, of new infection terms, and the *M* - matrix *V*, of the linear transition terms within the infected compartments of the model, are given, respectively, by:

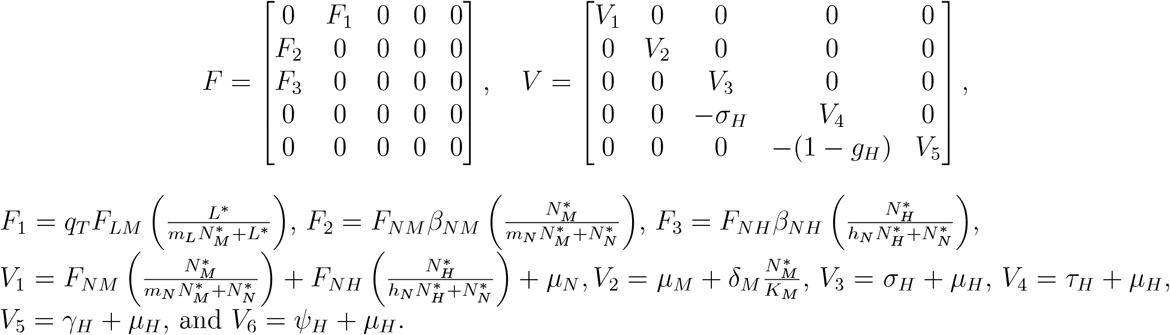

It is convenient to define the quantity (where *ρ* is the spectral radius):

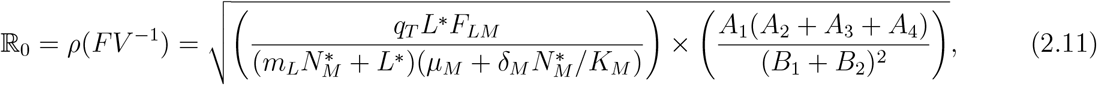

where: 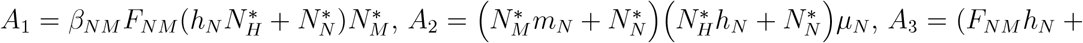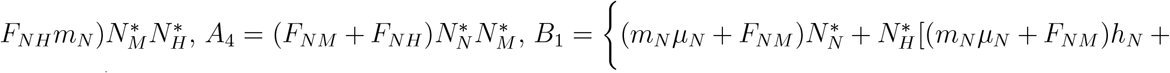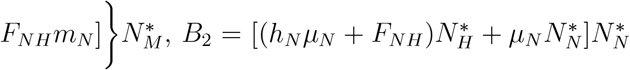. The result below follows from Theorem 2 of [106].

#### Theorem 2.2.

*Consider the model* (2.4)*–*(2.6) *with r*_0*M*_ *>* 1, Γ *>* 1, *T*_*A*_(*t*) *fixed at* 18°C, *and the values of* 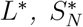, *and A*^*\**^, *as given in Tables* **??**. *The co-existence disease-free equilibrium of the model, given by* (2.9), *is locally-asymptotically stable whenever* ℝ_0_ *<* 1, *and unstable if* ℝ_0_ *>* 1.

The quantity ℝ_0_ is the *basic reproduction number* of the model {(2.4)–(2.6)} [106]. It measures the average number of new Lyme disease cases generated by a single infectious individual (or tick) if introduced in a wholly susceptible tick (or human) population. Theorem 2.2 says that a small influx of ticks or humans infected with Lyme disease will not generate a large outbreak in the community (harboring humans, ticks, mice, deer, and other reservoir hosts) if the basic reproduction number of the model is less than one. On the other hand, if ℝ_0_ *>* 1, this small influx will cause a large outbreak of Lyme disease in the human and other hosts populations in the community. In other words, Theorem 2.2 implies that Lyme disease can be effectively controlled or eliminated from the community, when the initial number of infected individuals or ticks is small enough (i.e., in the basin of attraction of the co-existence disease-free equilibrium), provided that the threshold quantity, ℝ_0_ can be brought to, and maintained at, a value less than one. Numerical simulations can be used to demonstrate that the solutions of the model converge to the co-existence disease-free equilibrium when ℝ_0_ *<* 1, and to an endemic equilibrium when ℝ_0_ *>* 1. On this basis, we propose the following conjecture.

#### Conjecture 2.3.

*The model* {(2.4)*–*(2.6)} *with r*_0*M*_ *>* 1, Γ *>* 1, *T*_*A*_(*t*) *fixed at* 18°C, *has at least one endemic equilibrium whenever* ℝ_0_ *>* 1. *This equilibrium is locally-asymptotically stable whenever it exists*.

Using the ranges of the estimated and fixed values of the parameters of the model, given in Tables **??** and **??**, respectively, and mean monthly temperature fixed at 18°C (corresponding to the tick season in Maryland, occurring during April–October), the value of the basic reproduction number (R_0_) for Lyme disease in Maryland ranges from 1.4 to 3.5, with a mean of 1.86. Thus, this study shows that, under the current ecological and environmental (local weather) conditions, Lyme disease will continue to persist in Maryland (since R_0_ *>* 1).

### 2.4 Model fitting and parameter estimation

The model {(2.4)–(2.6)} contains 26 parameters, and realistic values of 19 of these are available from the literature (see Table **??** of Supplementary Material **??**). The values of the remaining seven parameters lack empirical estimates from the literature and are instead determined by fitting the model to observed data. Specifically, the values of the parameters related to disease transmission from nymphs to humans (*β*_*NH*_), progression from the exposed to the symptomatic class (*σ*_*H*_), proportion of humans hospitalized with Lyme disease (*g*_*H*_), transition out of the symptomatic class (*τ*_*H*_), recovery rate (*γ*_*H*_), and rate of loss of infection-acquired immunity (*ψ*_*H*_), as well as the transmission rate parameter from nymphs to mice (*β*_*NM*_) will be estimated by fitting the model (using the values of the known parameters in Table **??**) with the relevant observed data (it should be mentioned that, for the fixed values of the model parameters in Table **??**, and the mean monthly temperature fixed at 18°, the threshold quantities *r*_0*M*_ and Γ for the state of Maryland take the values 24.61 and 872.74, respectively indicating the fact that current ecological conditions allow for the sustainability of the tick and mouse populations in the state of Maryland). Similarly, the values of *r*_0*M*_ and Γ for each of the 24 counties can be computed using the fixed mean monthly temperature specific to each county given in Table **??** of Supplementary Material **??** (for more details on the county-specific Lyme disease reported cases and mean monthly temperatures, see Figures **??** and **??** in Supplementary Material **??**). In particular, the model {(2.4)–(2.6)} is calibrated using cumulative Lyme disease case data reported by the CDC for each of the 24 counties in the State of Maryland, as well as for the entire state, over the period 2001 to 2022 [2]. The fitting is conducted using the model {(2.4)– (2.6)}, with the fixed monthly ambient temperature *T*_*A*_(*t*) for each county and for the entire state of Maryland as given in Table **??** of Supplementary Material **??**. The corresponding initial values of each of the state variables of the model used in the fitting procedure for each county and for the state of Maryland are presented in Section **??**. Standard least-squares regression method is used to minimize the sum of squared differences between the observed and the model-predicted cumulative number of Lyme disease cases for each county and for the entire state. A bootstrapping technique, with 10, 000 resamples, is used to estimate the respective 95% confidence interval for each fitted estimated parameter. The results of the fitting obtained, for each of the 24 counties, are depicted in Figure 5, and the values of the estimated parameters obtained from the fitting, together with their associated 95% confidence intervals, are tabulated in Table **??**. Figure 5 shows that the model fits the data reasonably well for each of the 24 counties (with *R*^2^ values ranging from 0.89 to 0.99, confirming the accuracy of the goodness of fit for each county). Similar goodness of fit is obtained for the entire state (see Figure **??** in Supplementary Material **??**).

**Figure 5:**
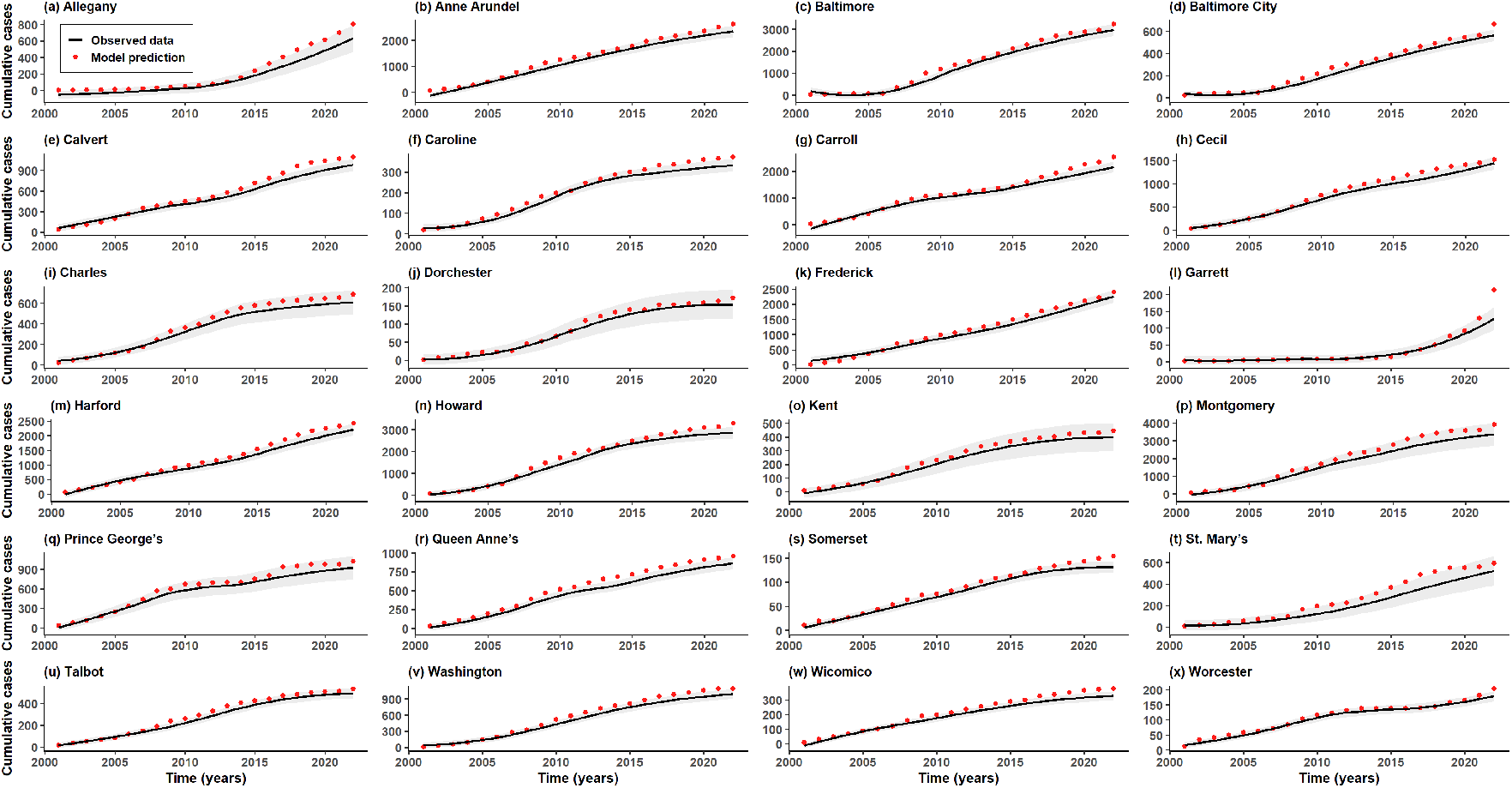
Data fitting of the model {(2.4)–(2.6)} to the yearly reported cumulative Lyme disease case data for each of the 24 counties in the state of Maryland for the time period from 2001 to 2022 (**a**–**x**). Mean monthly temperature value for each county during the tick season (tabulated in Table **??**) is used to evaluate the functional form of each of the 11 temperature-dependent parameters of the model (given in Section 2.2) for each county. The values of the fixed and fitted parameters, and their ranges (given in Tables **??** and **??**, respectively) are also used in the fitting (for these estimated and fitted values, the reproduction numbers for mice and ticks, *r*_0*M*_ *>* 1 and Γ *>* 1, respectively, exceed one). Furthermore, the equilibrium values for the tick population 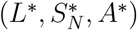 for each county, used in the data fitting, are given in Table **??** of Supplementary Material **??**. The shaded light gray regions in each of the 24 subplots represent the 95% confidence intervals.

### 2.5 Computation of optimal temperature range for ticks and mice abundance and Lyme disease transmission in the state of Maryland

In this section, the model {(2.4)–(2.6)} was simulated to determine the optimal temperature ranges for maximum population abundance of ticks and mice (as measured in terms of their respective reproduction numbers, Γ(*T*_*A*_) for ticks and *r*_0*M*_ (*T*_*A*_) for mice), as well as Lyme disease cases in humans (as measured in terms of the basic reproduction number, ℝ_0_(*T*_*A*_)), for the entire state of Maryland. These simulations are carried out using the typical temperature range during the ticks season in Maryland [104, 105, 108–110]. The results obtained, depicted in Figure 6, show significant increases in the value of each of the three reproduction numbers with increasing mean daily temperature (with maximum disease intensity and ticks and mice abundance at temperature values in the range *T*_*A*_ ∈ [17.0 − 20.5]°C), until a peak is reached at around 18.5°C, above which the values of each of the reproduction numbers decline markedly. The three metrics (Lyme disease cases/burden, ticks, and mice abundance) all peak at around the same temperature of 18.5°C in Maryland. In particular, Figure 6(**a**) shows that maximum abundance of ticks (i.e., tick survival, development, and host-seeking behavior) is attained in Maryland near this optimal temperature range. Similarly, Figure 6(**b**) shows that mice abundance (hence, their ability to transmit Lyme disease to the ticks) is maximized at this temperature range. Finally, Figure 6(**c**) shows that the state of Maryland experiences maximum Lyme disease transmission when the mean daily temperature is around 18.5°C. Thus, in summary, this study shows that Lyme disease activity in Maryland is maximized during time periods when the mean daily temperature lies within the optimal range 17.0−20.5°C, with peak intensity and ticks-mice abundance occurring at 18.5°C, suggesting that control efforts against ticks and their reservoir hosts should be intensified in Maryland during the time periods when the mean daily temperature values lie within the optimal range of 17.0 − 20.5°C.

**Figure 6:**
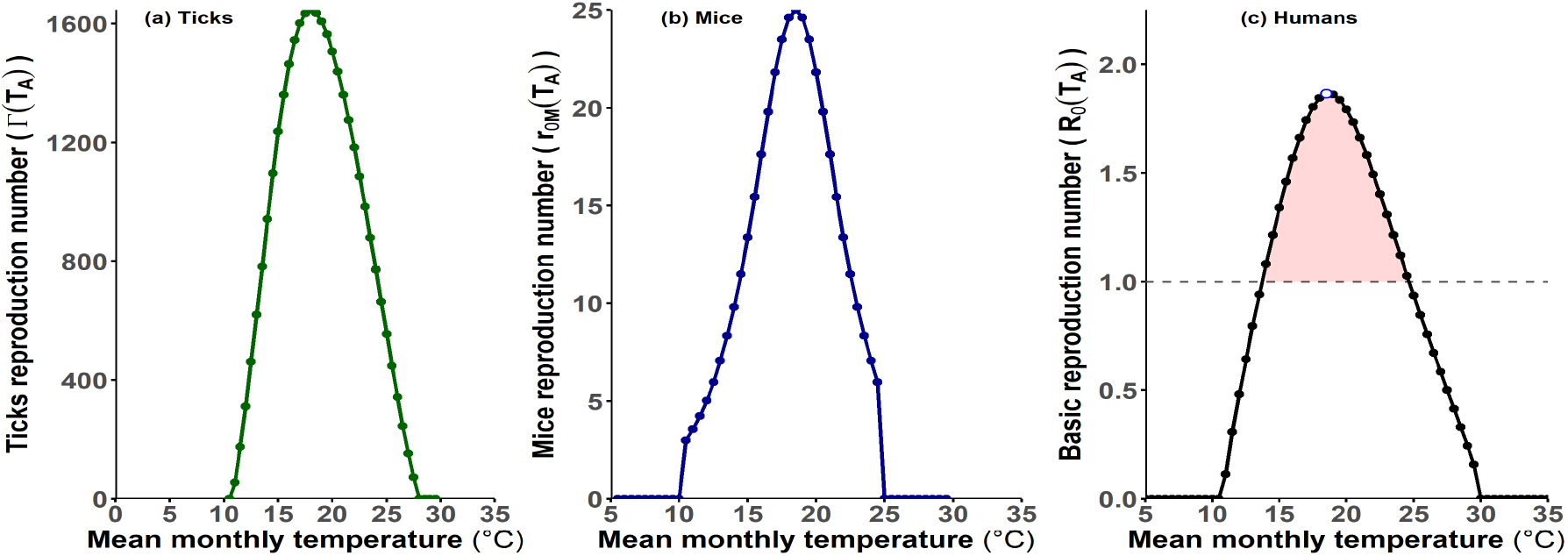
Simulations of the model {(2.4)–(2.6)} to assess the impact of temperature on Lyme disease burden and the population abundance of ticks and reservoir hosts (mice) in Maryland. (**a**) Effect of temperature on the ticks reproduction number (Γ(*T*)). (**b**) Effect of temperature on the mice reproduction number (*r*_0*M*_ (*T*)), and (**c**) Effect of temperature on the basic reproduction number (R_0_(*T*)). The values of the fixed and fitted (estimated) parameters of the model, used to generate these curves, are given in Tables **??** and **??** of Supplementary Material **??**, respectively. The equilibrium values for ticks population, 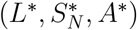 for the entire state of Maryland, are given in Table **??**. The brown-shaded region in (**c**) highlights the epidemic-prone regime where ℝ_0_ *>* 1, indicating sus-tained transmission potential.

The results presented in Figure 6 are consistent with those reported in [46, 105, 108, 110, 111], which also show that the activity, questing behavior, and survival of *I. scapularis* are strongly temperature-dependent, with optimal development and host-seeking observed in temperate conditions around 17.0–20.5°C [110]. Our findings highlight the importance of incorporating seasonal thermal suitability (for ticks and reservoir hosts) in Lyme disease risk assessments and control planning in Maryland and other temperate regions experiencing high Lyme disease prevalence. Additional simulations were carried out to predict the optimal temperature ranges for the maximum abundance of ticks and mice, as well as Lyme disease in humans, under the projected global warming scenarios of +2.5°C and +4.5°C increases in mean monthly global temperature. The results obtained, depicted in Figure **??** of Supplementary Material **??**, show that projected warming shifts the optimal temperature range for peak Lyme disease transmission and tick–mice abundance downward—from around 18.5°C under current conditions to approximately 16°C and 14.5°C under +2.5°C and +4.5°C scenarios, respectively. These decreases in the optimal range for maximum tick activity (and consequently Lyme disease intensity) reflect physiological responses of *I. scapularis* to warming—particularly thermal sensitivity of development, questing activity, and survival—where higher baseline temperatures cause heat stress that lowers the thresholds for peak tick activity and Lyme disease transmission [110, 112, 113]. The reduction in the optimal range is also associated with an overall reduction in the values of the control reproduction numbers, ℝ_0_(*T*), Γ(*T*), and *r*_0*M*_ (*T*). Thus, under projected global warming, Maryland may experience reduced Lyme disease risk in currently endemic zones, with a potential shift in suitability toward cooler regions. These results highlight the ecologically constrained effects of temperature on Lyme disease dynamics and the need for climate-informed, region-specific public health interventions.

#### Model-generated heat maps for Lyme disease burden in Maryland

In this section, the model {(2.4)–(2.6)} will be simulated, using the baseline values of the fixed and estimated parameters (given in Tables **??** and **??**) and the relevant demographic and mean monthly data during the optimal tick activity period for each of the 24 counties of Maryland (given in Table **??**), to generate a geospatial heat map for the cumulative number of Lyme disease cases in the state of Maryland. The simulation results depicted in Figure 7(**a**) show that, under the cur-rent global warming scenario, counties in Central Maryland (such as Montgomery, Baltimore, and Howard counties), but with exception of Baltimore city, are the hotspots for high Lyme disease burden in Maryland (Baltimore City experiences low to moderate Lyme disease burden, due, perhaps, to the reduced abundance and suitability of habitats for ticks and deer in the county [114]). Furthermore, counties in the Eastern (such as Cecil, Kent, and Worcester) and Western (such as Garrett and Allegany) Shore regions, as well as counties in Southern region of Maryland experience low to moderate Lyme disease burden—except Frederick county, which appears to be a high burden county, likely due to its proximity to hotspot counties in Central Maryland. Similarly, counties in the Southern region (i.e., Calvert, Charles, and St. Mary’s counties) experience low to moderate Lyme disease burden, likely due to favorable ecological and habitat suitability factors. Thus, these simulations show that, under the current global warming scenario, Lyme disease burden in Maryland is predominantly concentrated in the Central counties (which serve as the primary hotspot), while counties in the Western, Eastern, and Southern regions generally experience low to moderate Lyme disease burden, with the exception of Frederick county in the Western region and Cecil county in the Eastern region, which experience relatively high burden (similar geospatial maps are generated using the basic reproduction number, ℝ_0_, as a metric for Lyme disease burden across Maryland, and the results obtained are depicted in Figure **??** of Supplementary Material **??**). The model-generated geospatial map (depicted in Figure 7(**a**)) matches the observed data for the cumulative number of Lyme disease cases in the state of Maryland provided by the CDC [2] (depicted in Figure 7(**b**).

**Figure 7:**
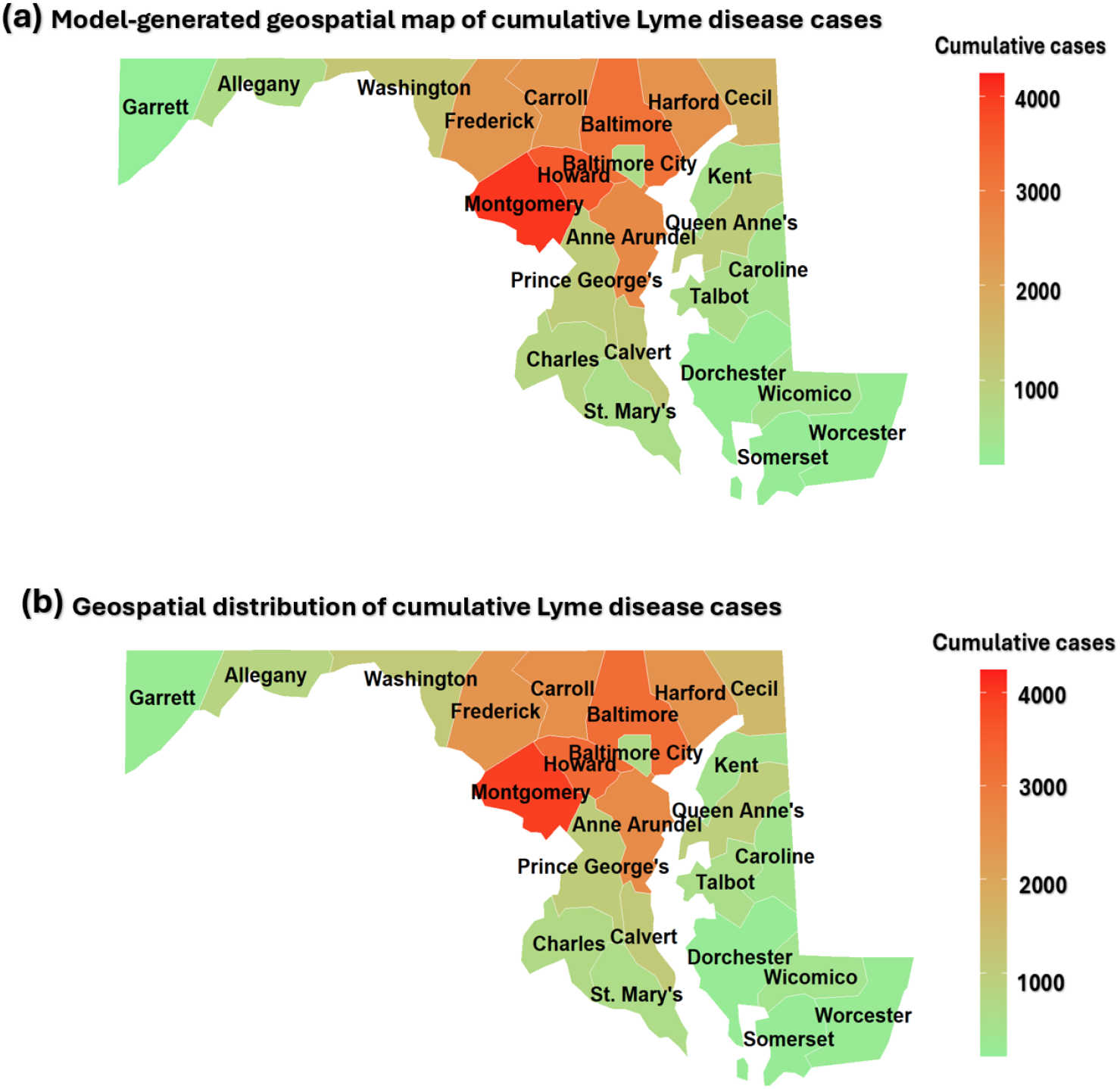
Geospatial map of the cumulative number of Lyme disease cases for the 24 counties of the state of Maryland for the period from 2001 to 2022. (**a**) Geospatial map generated using the model (2.4)–(2.6). (**b**) Geospatial map generated from the actual reported cumulative Lyme disease case data for Maryland for the same time period [2]. The model was fitted to the yearly reported Lyme disease case data to estimate the cumulative number of cases in each of the 24 counties. The mean monthly temperature values for each county during the tick season (tabulated in Table **??** of Supplementary Material **??**) were used to evaluate the functional forms of the eleven temperature-dependent parameters (see Section 2.2). The values of the fixed and fitted parameters, and their ranges (given in Tables **??** and **??**, respectively), were also used in the fitting (for these estimated and fitted values, the reproduction numbers for mice and ticks, *r*_0*M*_ *>* 1 and Γ *>* 1, respectively, exceed one). Furthermore, the equilibrium values for the tick population 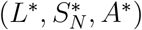 for Maryland,used in the data fitting, are given in Table **??. Risk scaling:** In this figure, county-level Lyme disease burden is classified according to the following risk scaling (in terms of cumulative cases): low transmission risk (*<* 900 cases), moderate transmission risk (900−1500 cases), high transmission risk (1500−2500 cases), and hotspot (ultra-high) transmission risk (*>* 2500 cases).

### 2.7 Impact of projected warming on Lyme disease burden in Maryland

The model {(2.4)–(2.6)} is now simulated to predict the geospatial distribution of Lyme disease across Maryland under the projected RCP 8.5 (Representative Concentration Pathway 8.5) global warming scenario of the Intergovernmental Panel on Climate Change [115], which anticipates a 2.5°C to 4.5°C (and up to 6.5°C in high-latitude regions) increase in mean global monthly temperature (i.e., monthly averages of daily mean temperature values) by 2100, relative to the pre-industrial era [39, 115–120]. For these simulations, Lyme disease burden in the state of Maryland is measured in terms of changes in the value of the basic reproduction number (ℝ_0_) for each county. Simulating the model for the case where the global mean monthly temperature is increased by 2.5°C reveals a further increase in Lyme disease burden in Central Maryland, expanding its primary hotspot region status, and extending transmission risk westward. Specifically, compared to the geospatial ℝ_0_ map for current warming scenario shown in Figure **??** of Supplementary Material **??**), the simulations for the 2.5°C warming scenario depicted in Figure 8 shows increased Lyme disease burden in Central Maryland, clear expansion into Western counties (such as Garrett and Allegany), relatively stable moderate Lyme disease burden across most of the Eastern counties, and a noticeable decline in the Lyme disease burden in the Southern counties. Specifically, under this projected 2.5°C temperature increase, counties in Central Maryland experience more intense hotspot status, while counties in Western Maryland (which previously experienced low to moderate burden) transition to high Lyme disease burden status; however, Maryland experiences an overall partial reduction in Lyme disease burden. This predicted westward shift of Lyme disease burden in Maryland is driven by the fact that this warming scenario pushes the local mean monthly temperatures into the estimated optimal range of approximately 15.0°C to 17.5°C (for maximum Lyme disease transmission) for these counties. In contrast, counties in Southern Maryland (including Calvert, Charles, and St. Mary’s), which initially experienced intermediate low-to-moderate Lyme disease burden under the current warming scenario, will experience low Lyme disease burden under the projected 2.5°C increase in mean monthly temperature (this decline is due to the fact that the projected warming pushes the mean monthly temperatures above their optimal for counties within this region, in addition to potential reductions in human exposure during hotter periods. Counties in Eastern Maryland generally maintain their current status under this scenario (i.e., most counties in the Eastern region remain in the low-to-moderate burden category, with the exception of Cecil County, which continues to experience moderate to high burden of the disease). This relative stability is due to the fact that most Eastern counties currently experience mean monthly temperatures that already fall within or close to the optimal thermal range for tick activity and Lyme disease transmission. Therefore, the additional 2.5°C warming does not substantially alter their ecological suitability, resulting in only marginal changes in transmission potential. In summary, our simulations show that the 2.5°C warming scenario will cause a geographic expansion of Lyme disease risk from Central into Western Maryland, together with a reduction in disease burden in Southern Maryland, and continued moderate risk in parts of Eastern Maryland (particularly Cecil County). This pattern reflects a westward shift and geographic expansion in habitat suitability, driven by local temperature changes across the regions. Thus, this study predicts that the projected 2.5°C increase in mean monthly temperature will show a dramatic increase in Lyme disease in Western Maryland (a region that currently experiences low to moderate burden of the disease), while counties in Southern and Eastern Maryland (which are currently experiencing low to moderate to high Lyme disease burden) will see a marked decrease in Lyme disease burden. Overall, this moderate warming scenario results in a partial statewide reduction in Lyme disease burden; however, it causes regional increases in the west and central regions, while leading to marked decreases in other areas.

**Figure 8:**
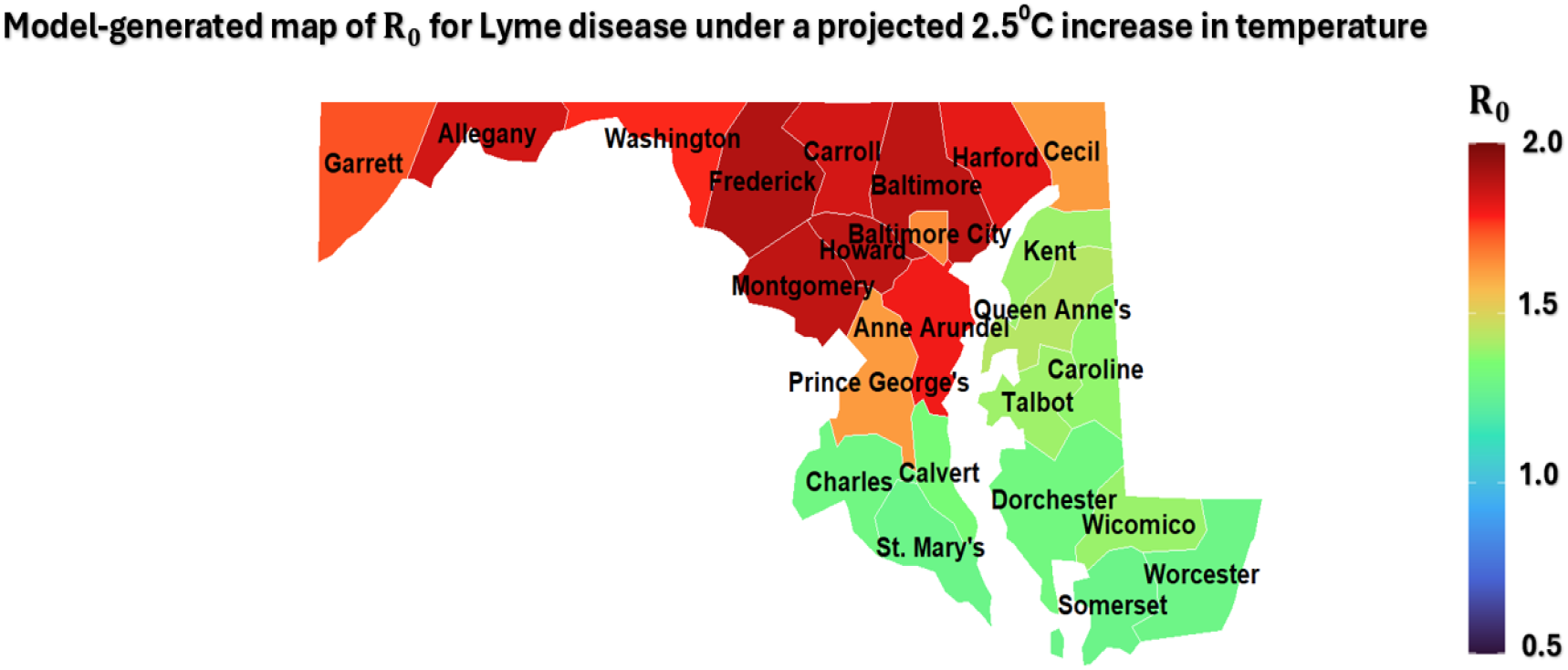
Simulation of the model {(2.4)–(2.6}) showing the geospatial distribution of the basic reproduction number (ℝ_0_(*T*_*A*_)) of the model across the 24 counties of Maryland for the projected global warming scenario with 2.5°C increase in mean monthly temperature (representing the lower bound projection under the RCP 8.5 climate scenario relative to the preindustrial levels). The ℝ_0_(*T*_*A*_) of Lyme disease was used to estimate the projected disease burden for each of the 24 counties under these warming conditions. The mean monthly temperature value for each county during the tick season (tabulated in Table **??**) is used to evaluate the functional form of each of the 11 temperature-dependent parameters of the model (defined in Section 2.2) for this scenario. The values of the fixed and fitted parameters, and their ranges (provided in Tables **??** and **??**, respectively), are also used in the simulations. For the fitted and estimated values, the reproduction numbers for mice and ticks satisfy *r*_0*M*_ *>* 1 and Γ *>* 1, respectively. The equilibrium values for the tick population 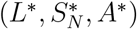 used in these projections are given in Table **??. Risk scaling:** In this figure, county-level Lyme disease transmission risk or potential is classified using the following scaling: low transmission risk (ℝ_0_ *<* 1.3), moderate transmission risk (1.3 ≤ R_0_ *<* 1.5), high transmission risk (1.5 ≤ R_0_ *<* 1.7), and hotspot (i.e., ultra-high) transmission risk (R_0_ − 1.7).

Similarly, the model {(2.4)–(2.6)} was further simulated under the high-end projection of a 4.5°C increase in mean monthly global temperature [115]. The results shown in Figure 9 reveal that counties in Western Maryland (e.g., Allegany and Garrett) will experience a marked increase in disease burden (i.e., their respective ℝ_0_ values will increase, and significantly exceed their corresponding values under the current warming scenario (Figure **??** of Supplementary Material **??**), making them new hotspots for Lyme disease in Maryland. In contrast, under this high warming scenario, counties in Central Maryland (e.g., Montgomery and Howard) will experience a decline in disease intensity, transitioning from hotspot zones to high-to-moderate burden zones. Eastern and Southern counties shift into low-transmission status, indicating decline in suitability for Lyme disease spread. In summary, moderate warming (+2.5°C) leads to a geographic expansion of Lyme disease burden—intensifying transmission in Central counties and elevating Western counties from low-to-moderate to high-burden status. However, more extreme warming (+4.5°C) drives a westward shift, with previously low-risk Western counties becoming new hotspots, while Central hotspots decline in intensity and Eastern and Southern counties transition to uniformly low-burden status.

**Figure 9:**
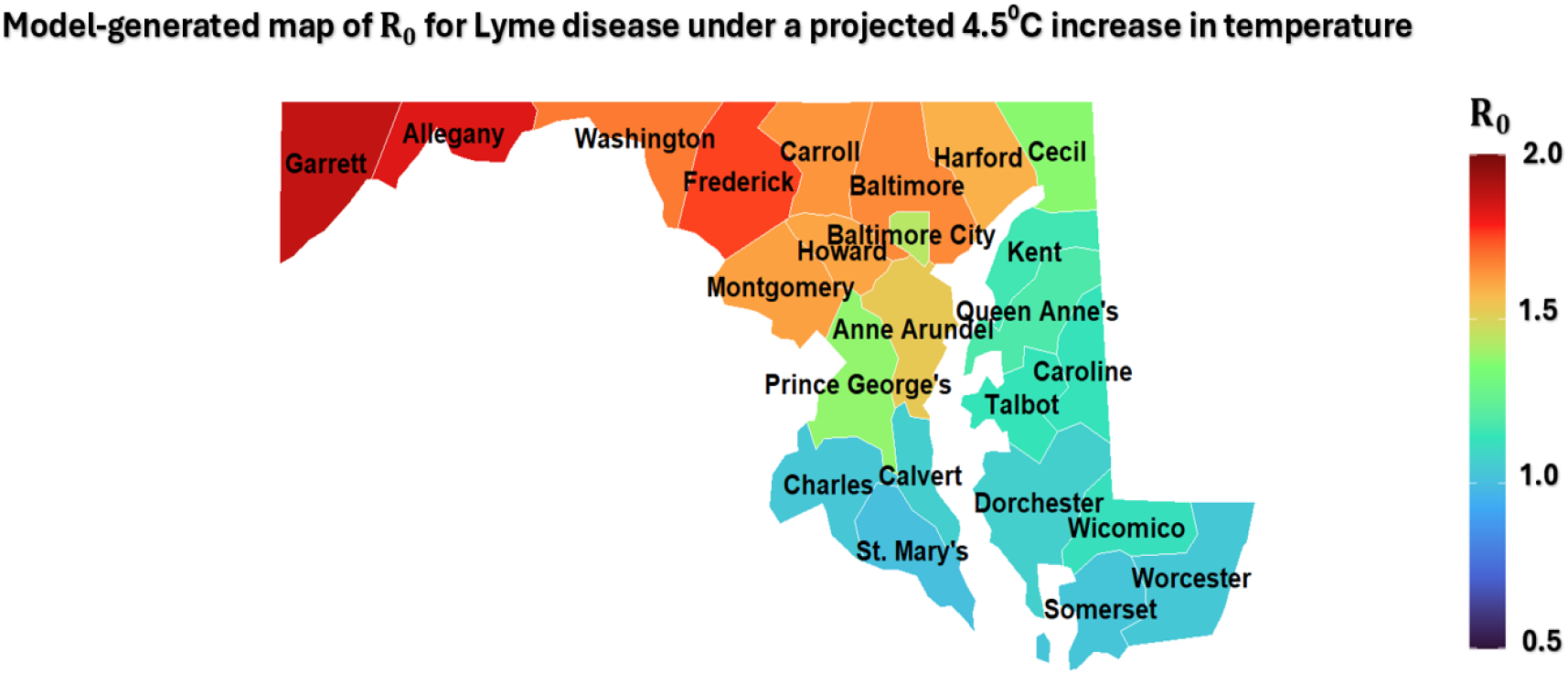
Simulations of the model {(2.4)–(2.6)} showing the geospatial distribution of the projected basic reproduction number (R_0_(*T*_*A*_)) of Lyme disease burden across the 24 counties of Maryland under a 4.5°C increase in mean monthly temperature—representing the upper bound projection under the RCP 8.5 climate scenario relative to preindustrial levels. The ℝ_0_(*T*_*A*_) was used to estimate the projected Lyme disease burden in each county under this elevated warming condition. County-specific mean monthly temperatures during the tick season (tabulated in Table **??**) were used to evaluate the functional forms of the eleven temperature-dependent parameters defined in Section 2.2. The fixed and fitted parameter values (and their ranges), provided in Tables **??** and **??**, were used in the simulations. The fitted parameters yield reproduction numbers for mice and ticks satisfying *r*_0*M*_ *>* 1 and Γ *>* 1, respectively. The corresponding equilibrium values for the tick population 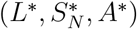 are listed in Table **??. Risk scaling:** In this figure, county-level Lyme disease transmission potential is classified using the following risk scaling: low (R_0_ *<* 1.3), moderate (1.3 ≤ ℝ_0_ *<* 1.5), high (1.5 ≤ ℝ_0_ *<* 1.7), and hotspot (ℝ_0_ − 1.7).

### 2.8 Sensitivity and uncertainty analyses

The model {(2.4)–(2.6)} contains numerous parameters. Hence, it is instructive to account for the impact of possible uncertainties in the estimates of the values of the parameters on the overall outcome of the numerical simulations of the model. This will be assessed using global uncertainty analysis, based on using Latin Hypercube Sampling (LHS) [121–123]. Furthermore, global sensitivity analysis will be carried out, using Partial Rank Correlation Coefficients (PRCCs), to determine the parameters that have the most influence on a chosen response function [121–123]. The LHS method, a stratified sampling without replacement technique, allows for the assessment of parameter variation across the full range of biological feasibility. In these analyses, which will be conducted for the entire state of Maryland, the basic reproduction number, R_0_, of the model is chosen as the response, and the baseline parameter values and ranges in Tables **??** and **??**, with ambient temperature fixed at 18°C and tick equilibrium values 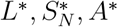 from Table **??**, will be used to generate the LHS samples. Each parameter of the model is assumed to follow a uniform distribution [122], and a total of 1,000 LHS samples—each representing one unique combination of all model parameters—were generated to evaluate variability in the response function, ℝ_0_.

The results obtained for the sensitivity analysis are depicted in Figure 10(**a**). This figure shows that the top PRCC-ranked parameters that are highly positively-correlated with the response function, ℝ_0_, are the transmission probability from infected mice to larvae (*q*_*T*_), larval feeding rate on mice (*F*_*LM*_), transmission rate from infected nymphs to mice (*β*_*NM*_), and nymphal feeding rate on mice (*F*_*NM*_). Similarly, the top PRCC-ranked parameters that are highly negatively-correlated with the response function are the density-dependent mortality rate of mice (*δ*_*M*_), mortality rate of nymphs (*µ*_*N*_), and the half-saturation constants for larval (*m*_*L*_) and nymphal (*m*_*N*_) feeding. Thus, these results suggest that public health intervention strategies that decrease (increase) the values of these top PRCC-ranked parameters will decrease the reproduction number (hence, reduce the burden of the disease in the community). In other words, this study shows that the following public health intervention strategies will decrease Lyme disease transmission and burden in the state of Maryland:

**Figure 10:**
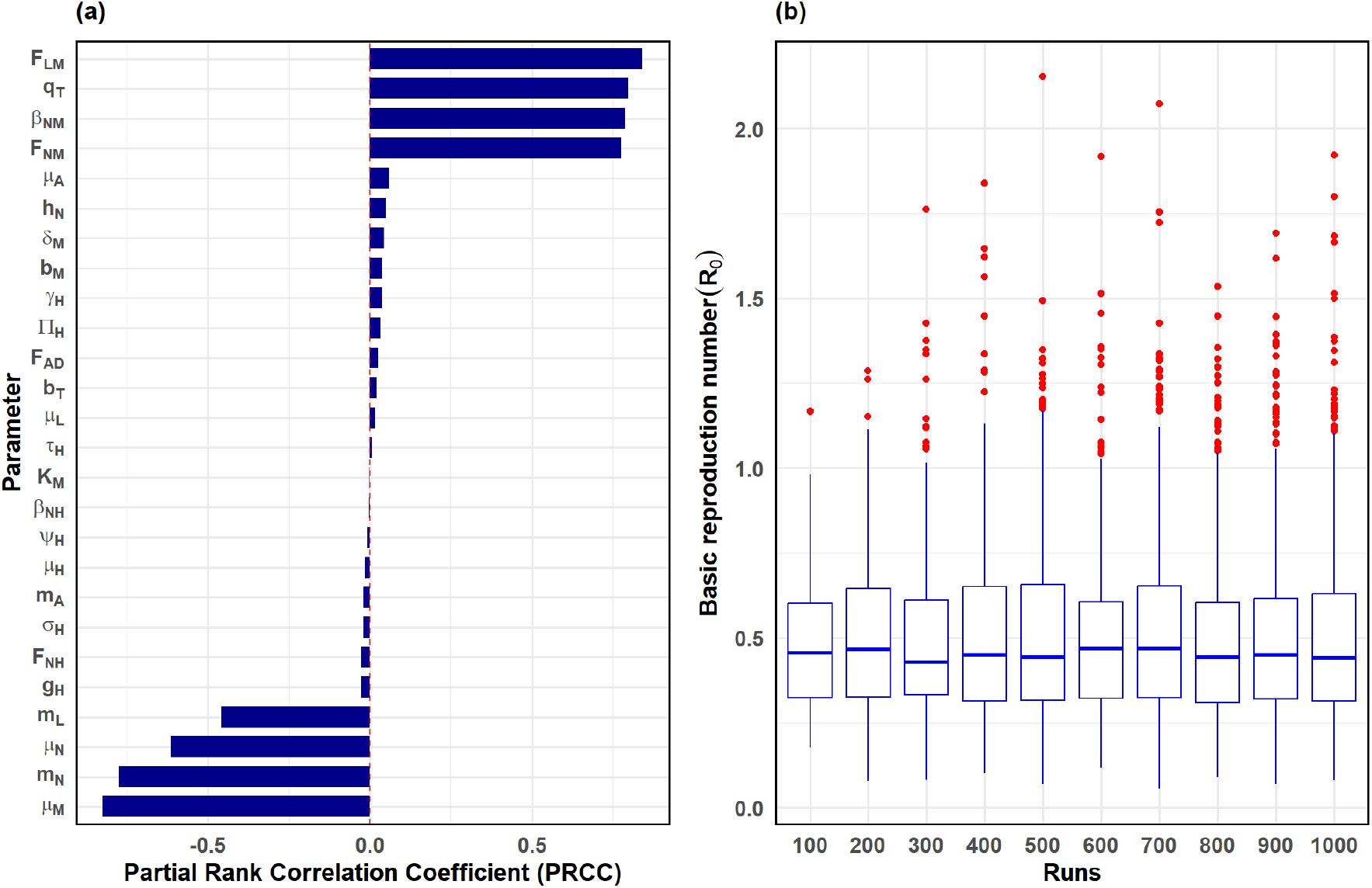
Simulations of the model {(2.4)–(2.6)} to assess the global sensitivity and uncertainty of the basic reproduction number (ℝ_0_) under a fixed temperature of *T* = 18°C, corresponding to the active tick season in Maryland. (**a**) Partial Rank Correlation Coefficients (PRCCs) quantifying the strength and direction of sensitivity of ℝ_0_ to key parameters. (**b**) Distribution of ℝ_0_ values from global uncertainty analysis using Latin Hypercube Sampling (LHS) across increasing sample sizes (“runs”) ranging from 100 to 1,000 parameter sets. Fixed and fitted parameter values and their ranges are provided in Tables **??** and **??**, respectively, so that *r*_0*M*_ *>* 1, and Γ *>* 1. The equilibrium values for ticks population 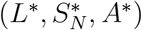 for Maryland are given in Table **??**.

i. Rodent-targeted bait treatment or population control campaigns, such as habitat modification or rodenticide, effectively reduce the density and recruitment of rodent hosts. These interventions primarily lower the larval and nymphal feeding rates on mice (*F*_*LM*_ and *F*_*NM*_) and reduce the force of infection to ticks by limiting the abundance of infected hosts, thereby decreasing the transmission probabilities (*q*_*T*_, *β*_*NM*_).
ii. Application of *acaricides*—a chemical vector control method—to rodent burrows, vegetation, or host animals effectively reduces the survival of immature and adult ticks. These control measures increase tick mortality rates (*µ*_*N*_ for nymphs and *µ*_*A*_ for adults), effectively disrupting the tick life cycle and suppressing ℝ_0_.
iii. Deployment of host-targeted tick control technologies, such as *permethrin*-treated bait boxes or tick tubes, interferes with tick-host interactions. These approaches increase the saturation effects (reflected by higher *m*_*L*_ and *m*_*N*_ values), thereby reducing tick feeding success on hosts and weakening transmission potential across both immature and adult stages.

Other strategies—such as rodent-targeted oral vaccines and landscape-level environmental management—offer complementary avenues for control. By combining ecological (habitat clearance-based) interventions with chemical (pesticide-based) methods (e.g., application of *acaricides* against ticks and host-targeted bait boxes), these integrated approaches can further disrupt transmission pathways and reduce the value of ℝ_0_, thereby significantly mitigating the burden of Lyme disease in Maryland and other endemic regions.

The results obtained for the uncertainty analysis, based on the 1, 000 samples taken for each parameter in the expression for R_0_, are depicted in the box plot in Figure 10(**b**). This plot shows that the value of R_0_ for the state of Maryland lie within the range [1.1, 2.125] with a mean of 1.58. Although the lower bound of this interval falls slightly below the epidemic threshold, the majority of sampled R_0_ values exceed 1, indicating a high likelihood of sustained transmission under current ecological and environmental conditions. To assess convergence of the uncertainty analysis, we conducted simulations using increasing sample sizes ranging from 100 to 1,000 (in increments of 100). The distribution of ℝ_0_ values stabilized beyond 800 samples, with minimal changes in both the mean and spread, thereby confirming the reliability of using 1,000 LHS samples for the final analysis.

Thus, in addition to showing that the mean value of ℝ_0_ for the state of Maryland is above one (suggesting that Lyme disease will continue to persist in the state under current ecological conditions and intervention coverages), this study identifies several parameters that have the highest impact on the trajectory and burden of the disease (as measured in terms of the value of the chosen response function, ℝ_0_). The consequence of this result is that the implementation of public health strategies that target these identified parameters (such as reducing rodent host density, disrupting larval and nymphal tick feeding, and applying acaricides—which collectively lower the key transmission pathways driving disease persistence) will lead to the effective control and mitigation of the burden of Lyme disease in the state of Maryland.

## 3 Numerical simulations of the model

The model {(2.4)–(2.6)} will now be adapted (extended) and simulated to assess the population-level impact of various control and mitigation measures against ticks, the reservoir hosts, and Lyme disease in Maryland. These simulations will be conducted at two values of the mean monthly temperature, corresponding to the onset (18°C) and the peak (20.5°C) of tick activity season in Maryland. The overall goal of these simulations is to evaluate the potential impact of the interventions on the geospatial dynamics of the *I. scapularis* population and the trajectory and burden of Lyme disease in the state of Maryland. The extensions and simulations are briefly described below.

### 3.1 Assessing the impact of control measures on Lyme disease dynamics

Although the model (2.4)–(2.6) does not explicitly account for intervention strategies against ticks, reservoir hosts, and/or Lyme disease, it can readily be extended to incorporate such strategies. Specifically, the model will be extended to include the following interventions:

a. **Habitat clearance (environmental management)**: This represents environmental management practices, such as vegetation trimming and leaf litter removal, to reduce tick habitats and larval exposure to infected mice [124, 125]. These actions target areas where ticks quest for blood meals from the hosts and breed, disrupting the enzootic cycle by limiting larval contact with reservoir hosts. This intervention can be accounted for in the model by rescaling the larval infection probability (*q*_*T*_), such that *q*_*T*_→ *q*_*T*_ (1− *ε*_*c*_), where 0 −*ε*_*c*_− 1 denotes the effectiveness of habitat clearance in reducing larval infection opportunities [125].
b. **Rodent-targeted control in residential communities**: This involves deploying *fipronil*-treated bait boxes to reduce tick populations on white-footed mouse (*P. leucopus*) in residential areas, schools, and playgrounds—locations where human exposure risk is elevated [95, 126]. The intervention works by killing attached ticks feeding on rodents, thereby disrupting the enzootic transmission cycle of *B. burgdorferi*. This control measure is incorporated into the model by rescaling the parameter for the rate of disease nymphs-to-mice transmission (*β*_*NM*_), such that *β*_*NM*_→ *β*_*NM*_ (1− *rε*_*r*_), where 0− *ε*_*r*_− 1 is the efficacy of the bait treatment in eliminating ticks, and 0 −*r* −1 represents the rodent bait coverage/proportion of the rodent population in the local environment that effectively encounters and interacts with the fipronil bait boxes (i.e., the proportion of the local rodent population exposed to the tick-killing treatment).
c. **Personal protection against tick bites**: This strategy consists of individual preventative measures, such as using insect repellents (e.g., DEET or *permethrin*) and wearing protective clothing, aimed at reducing the likelihood of nymphal ticks feeding on humans (the primary pathway for Lyme disease transmission in humans) [127, 128]. This strategy can be incorporated into the model by rescaling the parameter for the feeding rate of nymphs on humans (*F*_*NH*_), such that *F*_*NH*_→ *F*_*NH*_ (1 −*C*_*p*_*ε*_*p*_), where 0− *ε*_*p*_ −is the efficacy of repellents and 0 −*C*_*p*_− denotes the proportion of individuals complying with these measures. Studies indicate that DEET and *permethrin* can achieve up to 90% efficacy (of preventing human exposure to nymphal ticks) when applied properly [127].

The control reproduction number of the extended model, obtained by replacing the parameters *q*_*T*_, *β*_*NM*_, and *F*_*NH*_ in the expression for the basic reproduction number ℝ_0_ of the model given in Equation (2.11), by *q*_*T*_ (1 − *ε*_*c*_), *β*_*NM*_ (1 − *rε*_*r*_), and *F*_*NH*_(1 − *C*_*p*_*ε*_*p*_), respectively, is given by:

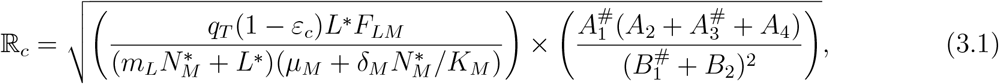

where the quantities *A*_2_, *A*_4_, *B*_2_ are as defined in Equation (2.11) and

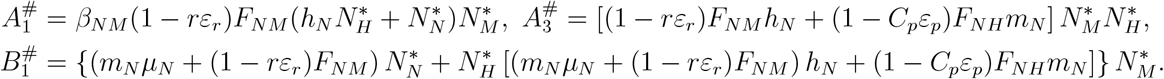

The extended model, given by {(2.4)–(2.6)} with the intervention-adjusted parameters, is now simulated to assess the population-level effectiveness of the interventions described above (and their combinations) in reducing the population abundance of ticks and reservoir hosts, and, consequently, on reducing Lyme disease burden in humans across the state of Maryland. The simulations, which are focused on evaluating the impact of the interventions on the value of the control reproduction number (ℝ_*c*_), will be carried out for two representative mean monthly temperature values, namely *T*_*A*_(*t*) = 18°C (baseline scenario, consistent with RCP 4.5) and *T*_*A*_(*t*) = 20.5°C (projected warming scenario, consistent with RCP 8.5) [115, 116]. These two temperature scenarios enable us to examine how global warming potential alters the effectiveness of interventions and the feasibility of Lyme disease elimination in the state of Maryland (to be potentially achieved by bringing and maintaining R_*c*_ to a value less than one (i.e., ℝ_*c*_ *<* 1)) or close to one.

#### 3.1.1 Assessing the impact of rodent-targeted control as the sole intervention

In order to assess the impact of rodent-targeted intervention implemented as the sole public health strategy (where infected rodents are targeted using bait stations containing acaricide or oral vaccines to reduce their capacity to sustain or transmit Lyme disease pathogens) on the transmission dynamics and control of Lyme disease in Maryland, the model {(2.4)–(2.6)} is simulated for the special case in which the other interventions described in Section 3.1, namely environmental clearance and personal protection, are not implemented. That is, for the rodents-targeted strategy as the sole intervention, we simulate the special case of the model with *ε*_*c*_ = *ε*_*p*_ = *C*_*p*_ = 0). The results obtained for the mean monthly temperature fixed at 18°C are depicted in the contour plot of the control reproduction number of the model, denoted by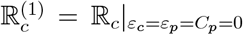, as a function of rodent bait coverage (*r*) and bait efficacy (*ε*_r_) in Figure 11(**a**). This figure shows that the control reproduction number 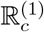 decreases with increasing efficacy or coverage of the rodent-targeted intervention. Specifically, the use of the rodent-based intervention as the sole control strategy can lead to the elimination of Lyme disease in Maryland if a bait treatment with efficacy of at least 75% (most standard rodent baits have efficacy in the range 70%–90%) [129, 130] is used and the coverage in its usage is high enough (e.g., ≥ 75%). For instance, setting bait efficacy to 75% (i.e., *ε*_*r*_ = 0.75) and bait coverage to 75% (i.e., *r* = 0.75), for temperature fixed at 18°C, gives 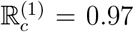 signifying the possibility of Lyme disease elimination in Maryland (in line with Theorem 2.2). For moderate levels of this intervention, such as the case when the bait coverage is set at 50%, which is more realistically attainable (while maintaining the bait efficacy at 75%), the control reproduction number increases to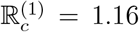,indicating that, although it reduces the disease burden, this (moderate) level of the rodent-targeted intervention is insufficient to eliminate Lyme disease in Maryland under the current (mean monthly temperature) climatic conditions. Similar results are obtained for the case with the case where the mean monthly temperature is set at *T*_*A*_ = 20.5°C. In this case, the value of the control reproduction number corresponding to the high and moderate coverage levels of this intervention are 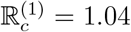 and 1.25, respectively (showing that, for this projected temperature value, neither the high nor the moderate level of this intervention can lead to Lyme disease elimination). Elimination is, however, feasible in this case if the coverage is at higher level (e.g., bait coverage *>* 80%; as illustrated in Figure 11(b)), which may not be attainable in practice. In summary, this study shows that, for the current mean monthly temperature in Maryland (18°C) and based on the parameter values used in our simulations (given in Tables **??** and **??**), the use of rodent-targeted control measures as a sole intervention (and with its efficacy fixed at 75%) will fail to eliminate Lyme disease in the state if the coverage in its usage is at low or moderate level (e.g., the bait coverage less than or equal to 50%). Elimination is, however, feasible if the coverage is high (e.g., −75%). For the case where the mean monthly temperature increased to 20.5°C (due to projected global warming), the rodents-targeted intervention (with its efficacy fixed at 75%) can only lead to disease elimination if the coverage level exceeds 80% (which may not be realistically attainable). Thus, this study further shows that increased temperature (due to global warming) makes the effort to effectively control or eliminate Lyme disease in Maryland using the rodents-targeted intervention (as a sole Lyme disease control strategy) more difficult, and suggest complementing the rodents-targeted strategy with other Lyme disease interventions, such as environmental clearance or personal protection, as explored below.

**Figure 11:**
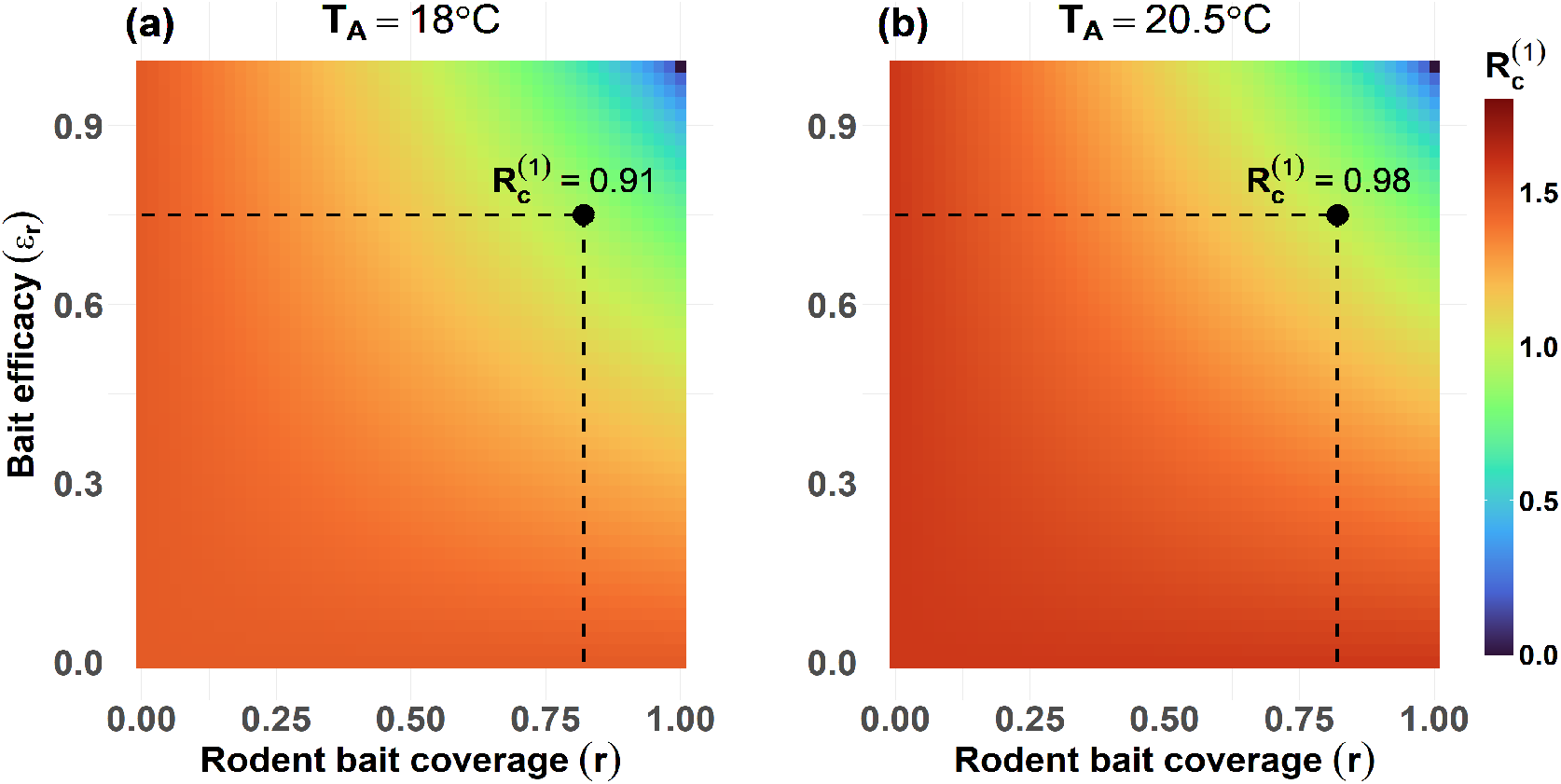
Effect of rodent-targeted strategy as a sole Lyme disease intervention in Maryland. Simulations of the model (2.4)–(2.6) showing contour plots of the control reproduction number 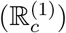 as a function of rodent treatment coverage (*r*) and bait treatment efficacy (*ε*_*r*_), for (**a**) mean monthly temperature fixed at *T* = 18°C (baseline condition) and (**b**) mean temperature fixed at *T* = 20.5°C (projected increase in climate change scenario). Other control parameters are held at baseline: personal protection compliance (*C*_*p*_ = 0), personal protection efficacy (*ε*_*p*_ = 0), and environmental control efficacy (*ε*_*c*_ = 0). Fixed and fitted parameter values and their ranges are provided in Tables **??** and **??**, respectively, so that *r*_0*M*_ *>* 1, and Γ *>* 1. The equilibrium values for ticks population 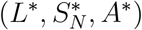 for Maryland are given in Table **??**. The intersections of the dashed vertical and horizontal lines represent the value of 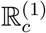 for the case where (*r, ε*_*r*_) = (0.75, 0.8).

#### 3.1.2 Assessing the combined impact of rodent baiting and habitat clearance measures

To evaluate the combined impact of rodent-targeted and habitat clearance interventions—implemented as a dual public health strategy (where infected rodents are targeted using bait stations containing acaricide or oral vaccines, and habitat clearance reduces tick populations by modifying their environment)—on the transmission dynamics and control of Lyme disease in Maryland, the model {(2.4)–(2.6)} is simulated for the special case in which personal protection is not implemented (i.e., *C*_*p*_ = *ε*_*p*_ = 0). Here, too, the rodent bait efficacy is fixed at 75% (*ε*_*r*_ = 0.75), in line with the reported standard effectiveness of most rodent baits [129, 130]. The control reproduction number associated with this simulation, denoted by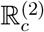, is given by 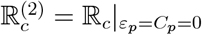 (with the bait efficacy maintained at 75%). The results obtained, for the mean monthly temperature fixed at 18°C, are depicted in the contour plot of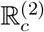, as a function of rodent bait coverage (*r*) and habitat clearance effectiveness *ε*_*c*_) in Figure 12(**a**). This figure shows that 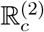 decreases with increasing coverage (*r*) and efficacy (*ε*_*r*_) of the rodent-targeted, as well as with increasing effectiveness of the habitat clearance intervention (*ε*_*c*_). Specifically, elimination can be achieved in Maryland by combining the rodent-based strategy at high efficacy with moderate levels of bait coverage and habitat clearance effectiveness. For instance, setting the bait efficacy at 75% (i.e., *ε*_*r*_ = 0.75), bait coverage and habitat clearance effectiveness at 50% each (i.e., *r* = *ε*_*c*_ = 0.50), and fixing the mean monthly temperature at 18°C, gives 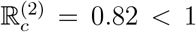, suggesting that disease elimination is feasible (in line with Theorem 2.2).

**Figure 12:**
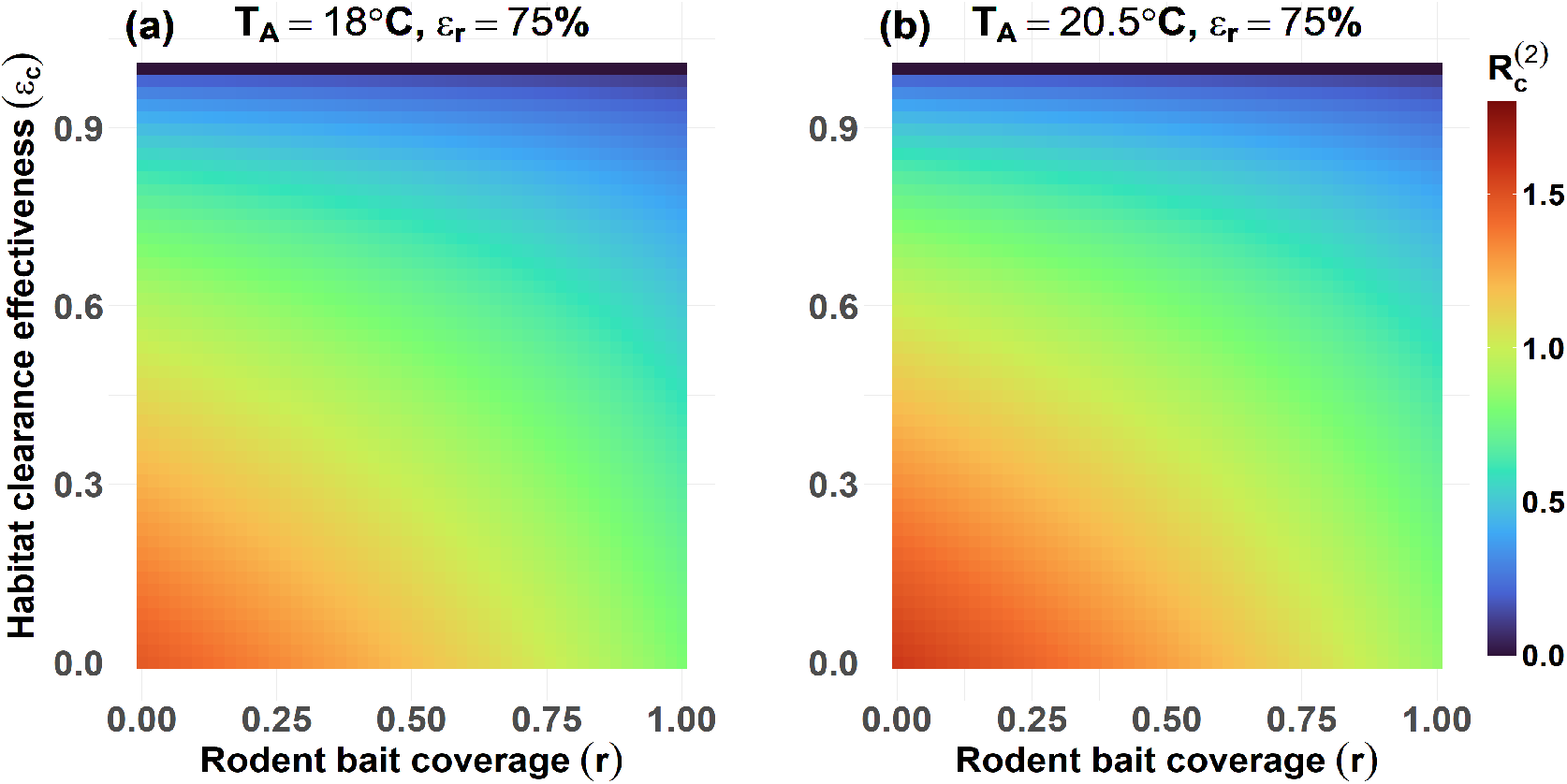
Effect of combined rodent-targeted and habitat clearance strategies. Simulations of the model (2.4)–(2.6) showing contour plots of the associated control reproduction number 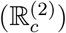,as a function of rodent treatment coverage (*r*) and habitat clearance effectiveness (*ε*_*c*_), for (**a**) mean monthly temperature fixed at *T*_*A*_ = 18°C (baseline condition) and (**b**) mean temperature fixed at *T*_*A*_ = 20.5°C (projected increase in climate change scenario). The rodent bait efficacy is fixed at *ε*_*r*_ = 0.75, and other control parameters are held at baseline: personal protection compliance (*C*_*p*_ = 0) and personal protection efficacy (*ε*_*p*_ = 0). Fixed and fitted parameter values and their ranges are provided in Tables **??** and **??**, respectively, so that *r*_0*M*_ *>* 1, and Γ *>* 1. The equilibrium values for the tick population 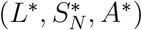 for Maryland are given in Table **??**.

Under this same temperature setting, increasing the bait coverage and clearance effectiveness to 75% (i.e., *r* = *ε*_*c*_ = 0.75), while maintaining the bait efficacy at 75%, reduces the reproduction number further to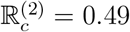, thereby strengthening reduction in disease burden and enhancing elimination prospect under the high effectiveness level of this combined intervention. The value of the control reproduction corresponding to the moderate level of this intervention (i.e., *ε*_*r*_ = 0.75, *r* = *ε*_*c*_ = 0.5) slightly increases to 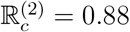 at the projected mean monthly temperature 20.5°C (also suggesting disease elimination; the number decreases to 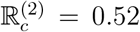 under the high effectiveness level, with *ε*_*r*_ = *r* = *ε*_*c*_ = 0.75). In summary, this study shows that, for the current mean monthly temperature (18°C), the combined rodent-targeted and habitat clearance strategy with high rodents bait efficacy (at least 75%) will lead to the elimination of Lyme disease in Maryland even with moderate (e.g., 50%) coverage (Figure 12**(a)**). This contrasts with the case of the rodents-targeted strategy as a sole intervention (discussed in Section 3.1.1), which failed to lead to elimination at moderate coverage levels. This combined strategy (with high bait efficacy) also leads to disease elimination for the projected mean monthly temperature (20.5°C) even with moderate coverage levels and habitat clearance effectiveness (Figure 12**(b)**), although such elimination requires a little more effort, since the values of the reproduction numbers are larger (despite being less than one) than for the corresponding case with mean monthly temperature fixed at 18°C. Overall, these simulations show that combining the rodents-based strategy with habitat clearance intervention (even at moderate coverage and habitat clearance effectiveness levels, but with high enough bait efficacy) significantly enhances the prospect of Lyme disease elimination in Maryland under the current and the projected mean monthly temperature regimes.

#### 3.1.3 Impact of hybrid rodent-targeted, personal protection, and habitat clearance

In this section, the model {(2.4)–(2.6)} is simulated to assess the population-level effectiveness of a hybrid strategy that combines the aforementioned three strategies (rodent baiting, personal protection, and habitat clearance) on the control of Lyme disease in Maryland. For these simulations, the following three effectiveness levels of the hybrid strategy are considered and compared against a worst-case scenario with no interventions implemented (i.e., *ϵ*_*c*_ = *ϵ*_*p*_ = *C*_*p*_ = *ϵ*_*r*_ = *r* = 0).

i. **Low effectiveness level:** This entails setting the habitat clearance effectiveness, personal protection efficacy and coverage, as well as bait coverage at 25% each (i.e., *ε*_*c*_ = *ε*_*p*_ = *C*_*p*_ = *r* = 0.25), while maintaining the rodent bait efficacy at a fixed value of 75% (i.e., *ε*_*r*_ = 0.75). For this effectiveness level, the associated control reproduction number is 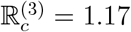 at the current mean monthly temperature (*T*_*A*_ = 18°C) and 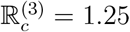 at the projected 20.5°C. Hence, since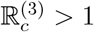, elimination is not feasible under this hybrid strategy at either temperature.
ii. **Moderate effectiveness level:** For this level, the habitat clearance effectiveness, personal protection efficacy and coverage, and bait coverage are all set at 50% (i.e., *ε*_*c*_ = *ε*_*p*_ = *C*_*p*_ = *r* = 0.5), with the bait efficacy fixed at *ε*_*r*_ = 0.75. Under these conditions, the control reproduction number takes the values 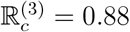 and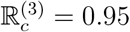.95 at 18°C and 20.5°C, respectively. Thus, elimination is feasible at both temperatures, although the margin is narrower under the projected warming scenario.
iii. **High effectiveness level:** In this case, habitat clearance effectiveness, personal protection efficacy and coverage, and bait coverage are all set at 75% (i.e., *ε*_*c*_ = *ε*_*p*_ = *C*_*p*_ = *r* = 0.75), while the rodent bait efficacy remains fixed at *ε*_*r*_ = 0.75. The corresponding control reproduction numbers are 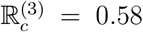 at 18°C and 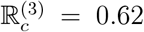 at 20.5°C, confirming that elimination is consistently achievable under this high-effectiveness strategy, even with projected climate warming.

The simulation results obtained, depicted in Figure 13, show reductions in the cumulative number of Lyme disease cases in humans, with increasing effectiveness level of the hybrid strategy, in comparison to the worst-case scenario for both the current and projected mean monthly temperature scenario. This reduction is particularly more pronounced for the case of the current mean monthly temperature (Figure 13(a)), compared to the case for the projected temperature (Figure 13(b)). The control reproduction numbers corresponding to the low, moderate, and high-effectiveness levels of this hybrid strategy are, respectively, 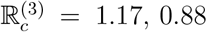, and 0.58 (it should be noted that the value of the control reproduction number corresponding to the worst-case scenario is 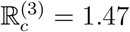, confirming that each of the three effectiveness levels of the hybrid strategy significantly reduces the disease burden, in comparison to the worst-case scenario). Thus, the hybrid strategy can lead to Lyme disease elimination in Maryland under the current temperature regime if implemented at the moderate or high effectiveness level described above. For the case of the projected mean monthly temperature, the value of the control reproduction number corresponding to the worst-case scenario is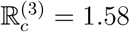, and the values corresponding to the low, moderate and high effectiveness levels of this hybrid strategy are 1.25, 0.95, and 0.62, respectively (showing, also, that elimination is feasible for moderate or high effectiveness levels). In summary, this study shows that the prospect of Lyme disease elimination in Maryland are promising using the hybrid strategy implemented at potentially attainable moderate level. This study highlights the importance of integrated multi-faceted strategies for the effective long-term control or elimination of Lyme disease in Maryland.

**Figure 13:**
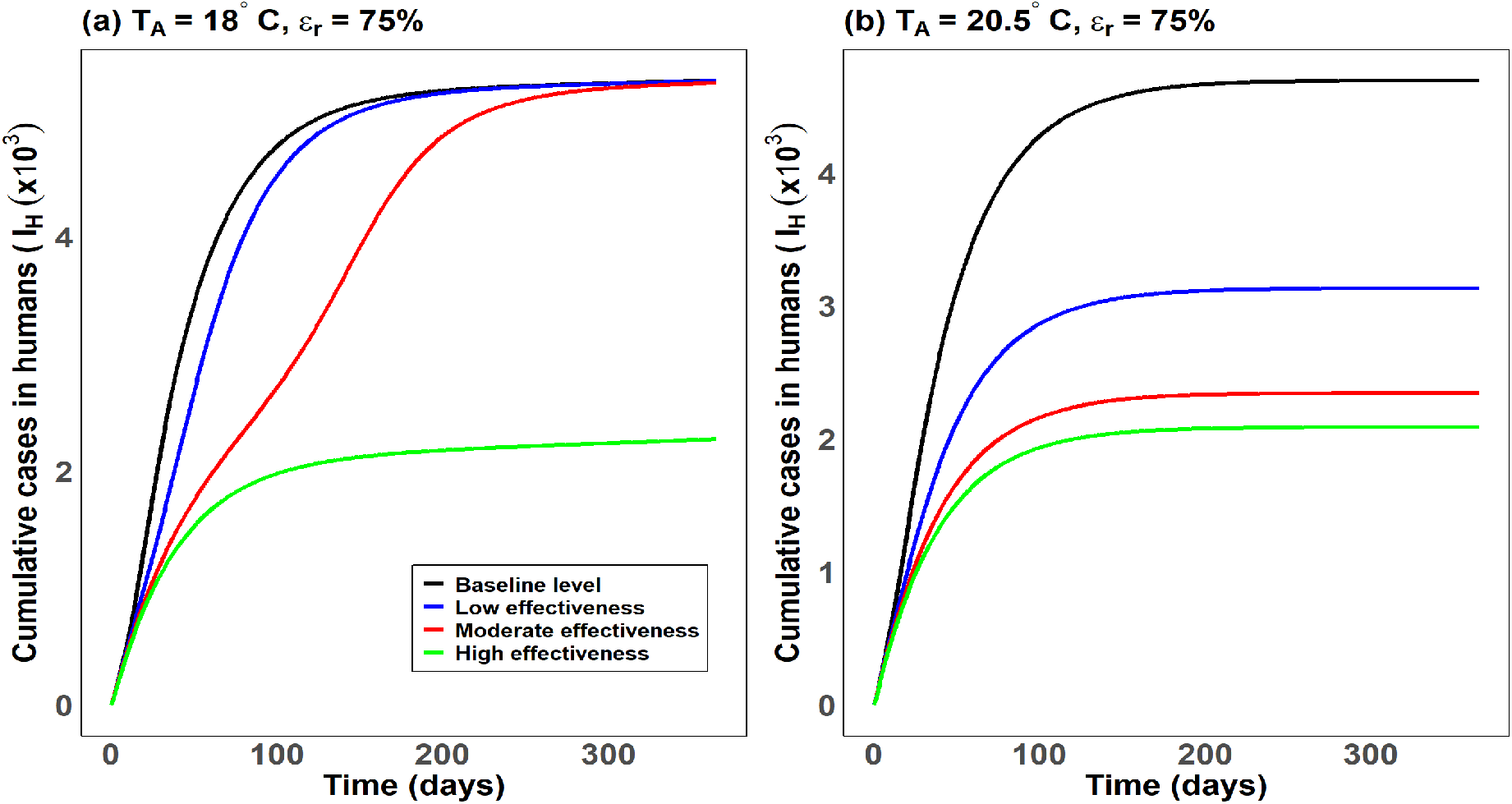
Simulation of the model (2.4)–(2.6) illustrating the impact of a hybrid intervention strategy—combining rodent baiting, personal protection, and habitat clearance—on cumulative human Lyme disease cases (*I*_*H*_). Panels (**a**) and (**b**) display the time series of cumulative new human infections under the baseline temperature (*T*_*A*_ = 18°C) and the projected elevated temperature (*T*_*A*_ = 20.5°C), respectively. In each panel: (i) the black curve denotes the no-intervention scenario (*ε*_*c*_ = *ε*_*p*_ = *C*_*p*_ = *r* = 0); (ii) the blue curve represents the low-effectiveness level (*ε*_*c*_ = *ε*_*p*_ = *C*_*p*_ = *r* = 0.25, with *ε*_*r*_ = 0.75); (iii) the red curve corresponds to the moderate-effectiveness level (*ε*_*c*_ = *ε*_*p*_ = *C*_*p*_ = *r* = 0.50, with *ε*_*r*_ = 0.75); and (iv) the green curve depicts the high-effectiveness level (*ε*_*c*_ = *ε*_*p*_ = *C*_*p*_ = *r* = 0.75, with *ε*_*r*_ = 0.75). Parameter values used in the simulations are listed in Tables **??** and **??**, under ecological conditions where *r*_0*M*_ *>* 1 and Γ *>* 1. Equilibrium values for the tick population 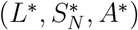 in Maryland are given in Table **??**.

## 4 Discussion and conclusion

Lyme disease, caused by *B. burgdorferi* and transmitted primarily by *I. scapularis* ticks and maintained in enzootic cycles involving white-footed mice (*P. leucopus*) and deer, is a rising public health threat in the state of Maryland, with cases increasing over the last three decades [2, 13]. We developed a mechanistic mathematical model, which explicitly accounts for temperature-dependent tick life stages, host–pathogen interactions, and climate change projections under two greenhouse emission scenarios (moderate and high emission scenarios, where the mean global monthly temperature is expected to increase by 2.5°C and 4.5°C, respectively, by 2100, relative to the pre-industrial era [116]. The objective was to quantify the impact of changes in temperature and ecological conditions on the abundance and distribution of ticks and Lyme disease burden across the 24 counties in the state of Maryland. The model, which specifically accounted for 11 temperature-dependent parameters for tick development, survival, feeding, and host reproduction (Figure 3), was calibrated and validated using Lyme disease and temperature data for the state of Maryland for the years 2001-2022 [2, 102].

Theoretical analysis of the calibrated model showed that the long-term dynamics of the disease is governed by the value of an epidemiological threshold, known as the basic reproduction number (denoted by R_0_), which governs the elimination of the disease when it is less than one, and persistence when it exceeds one. Using realistic set of parameter values (corresponding to the current ecological and environmental conditions in Maryland), it was shown that the basic reproduction number for Lyme disease in Maryland range from R_0_ = 1.1 to R_0_ = 1.86 (with a mean of R_0_ = 1.58). Hence, this study indicates that, under the present ecological and environmental conditions, Lyme disease will continue to persist in Maryland.

Our study showed, by simulating the calibrated model {(2.4)–(2.6)}, that under current ecological and environmental conditions in Maryland, tick activity (as well as reservoir host abundance) and Lyme disease transmission risk are maximized when the mean daily temperature during the tick season (April–October) lies within the optimal range of 17.0°C–20.5°C, with peak transmission attained at approximately 18.5°C. This optimal temperature range is typically recorded in Maryland during the active tick season, suggesting that control measures against tick exposure and reservoir host contact should be intensified during this period. The optimal temperature range generated from this study is consistent with other modeling and empirical studies of *I. scapularis* spread in North America, which also report maximal host-seeking (questing) activity and high tick survival when the mean daily temperature lies in the range 17.0–20.5°C [46, 105, 108, 110–112, 131]. For the projected moderate greenhouse emission scenario (RCP 4.5), where the mean global temperature is expected to increase by about 2.5°C by 2100 relative to pre-industrial levels, our study predicted that the optimal range for maximum tick activity and Lyme disease intensity in Maryland will decrease downward to approximately 15.0°C–17.5°C, with peak transmission occurring at around 16°C. This implies that moderate warming will move the optimal peak from 18.5°C to 16°C. Consequently, hotspot central counties (such as Montgomery, Howard, and Baltimore) and counties in southern and eastern Maryland (such as Calvert and Cecil), which are currently at or near the peak temperature (18.5°C), will move further away from the new (reduced) optimal temperature range, consequently resulting in a decline of Lyme disease intensity and burden. In contrast, cooler western regions (such as Garrett and Allegany), which currently record temperatures below the current optimal range (and experience very low Lyme disease burden), will warm into the new optimal temperature range of 16.0–17.5°C. Consequently, these counties will move closer to the peak of the new optimal temperature range, making them more vulnerable for increased tick activity and sustained transmission. Since most of the central and eastern counties that are experiencing temperatures above the optimal range, and predicted to experience lower Lyme disease burden, are very populated (accounting for about 70% of the population of Maryland), and the western counties that are warming to the peak of the new optimal temperature range for maximum Lyme disease burden are sparsely populated (accounting for about 30% of the total population of Maryland), the overall effect of the moderate emission scenario is a net reduction in Lyme disease burden cases across Maryland. Similarly, under the high-emission scenario (RCP 8.5), with the mean global temperature projected to increase by about 4.5°C by 2100, the optimal temperature range for maximum tick activity and Lyme disease intensity decreases further to 13.5°C–15.5°C, with peak transmission occurring at around 14.5°C. This represents a more significant reduction in the optimal temperature range, and counties in central, southern, and eastern Maryland will be recording much warmer temperatures (around 22°C) far above the new optimal temperature range (and much hotter for tick’s activities and survival). For this high emission scenario, the cooler western counties will mostly fall fully within this new optimal temperature range, making them more vulnerable for tick activities and potentially emerge as new hotspot for Lyme disease transmission. However, since these western counties are sparsely populated, compared to the central and eastern regions, the overall effect of the high emission scenario is an even greater statewide reduction in Lyme disease burden, compared to the moderate emission scenario. In summary, our results indicate that global warming, under both moderate (RCP 4.5) and high (RCP 8.5) emission scenarios, will push many hotspot and moderate Lyme disease counties in central, southern, and eastern Maryland (which constitute over 70% of the state’s population) above the optimal temperature range, resulting in reduced transmission by 2100. In contrast, western counties (accounting for about 30% of the population) will move closer to, or reach, the peak of the new optimal range and will, consequently, see increased Lyme disease burden by 2100. These changes in the optimal temperature range for maximum *I. scapularis* activity in Maryland reflect the tick’s physiological responses to warming—particularly the thermal sensitivity of tick’s development, questing activity, and survival (with rising baseline temperatures cause the physiological optimum of the tick to decrease toward cooler temperature ranges due to heat stress for temperatures above the optimal range [104, 110, 112, 113]).

Geospatial projections under the current ecological and environmental conditions accurately reproduced observed patterns, with Central Maryland—especially Montgomery, Baltimore, and Howard counties—emerging as consistent hotspots for Lyme disease transmission. Moderate warming under RCP 4.5 is projected to intensify transmission in these central counties and expand risk westward into cooler areas such as Garrett County, where warming is projected to push temperatures within the optimal range for maximum Lyme disease transmission intensity. However, most counties in the southern and eastern regions are projected to experience reduced Lyme disease burden because warming pushed them outside the optimal range (where *I. scapularis* activity, development, and survival, etc., are greatly reduced). Under the high-emission RCP 8.5 scenario, Lyme disease transmission risk shifts markedly, where current hotspots in central Maryland are projected to see more declining Lyme disease transmission risk and burden (due to thermal overshoot), while western counties, such as Garrett and Allegany, will become new primary hotspots. Similarly, the southern and eastern counties experience further reductions in Lyme disease burden due to the reduced suitability of environmental conditions for the tick vector as temperatures exceed the optimal range. These results indicate that moderate warming will expand risk into central and western Maryland despite an overall statewide reduction, whereas extreme warming will drive a more pronounced westward shift in Lyme disease burden—highlighting the need for geographically adaptive public health strategies. The model developed in this study was simulated to assess the population-level impact of various intervention strategies, notably rodent baiting, habitat clearance, and personal protection against exposure to ticks. The results obtained showed that, under current climatic conditions (with mean daily temperature during the tick season fixed at 18°C), rodent-targeted control alone can substantially reduce Lyme disease burden, with elimination attainable in Maryland if the bait efficacy and coverage are high enough (at least 75% each). Under projected moderate warming in the RCP 4.5 scenario (where the mean mean daily temperature during the tick season is projected to be 20.5°C), elimination using rodents-targeted control alone remains possible (but requires coverage levels exceeding 80% if bait control with efficacy 75% is used). This is because of the increased tick–host activity at (increased) warmer temperatures. Combining rodent baiting with habitat clearance makes elimination feasible at more moderate and attainable coverage levels (e.g., 50% bait and personal protection coverages, with bait efficacy at 75%) under both the moderate and high emission scenarios. We showed that a hybrid strategy, which combines all three interventions, is the most effective, as expected. At moderate level of implementation of this strategy (i.e., 75% bait efficacy combined with 50% coverage each for habitat clearance, bait, and personal protection), elimination is achievable under both emission scenarios (with elimination achieved faster if coverages are higher than 50%). Under the two warming scenarios, Lyme disease elimination requires higher effectiveness and coverage levels of control intervention efforts—such as higher rodent bait coverage and more effective habitat clearance—highlighting the need to scale control strategies to future climate conditions. Overall, these findings emphasize that integrated, climate-adaptive control strategies offer the most reliable pathway for sustained control of the burden and potential elimination of Lyme disease in Maryland. Our results are consistent with previous studies showing that rodent-targeted interventions can be highly effective when implemented at sufficiently high coverage [29, 132, 133]. The enhanced effectiveness of the combined or hybrid strategy also aligns with evidence from other vector-borne disease systems, where integrated approaches that target multiple pathways outperform single interventions [82, 134]. Thus, while climate change is expected to raise the thresholds of intervention efforts for Lyme disease elimination, our study demonstrates that a hybrid strategy that combines rodent-targeted, habitat clearance, and personal protection strategies remains a robust and evidence-supported pathway for long-term Lyme disease control and possible elimination in Maryland.

While our mechanistic modeling study offers a strong, data-driven framework for assessing the population-level impact of climate change on the geospatial distribution and burden of Lyme disease in Maryland, certain aspects remain beyond its current scope and present opportunities for improvement. For example, although the model’s assumption of homogeneous risk exposure and intervention uptake within each county enables clear, large-scale insights, it does not account for the local heterogeneity in land use, tick habitat, host biodiversity, or human behavior. Likewise, the current study focused on temperature as the primary climatic driver, without accounting for co-infections with other tick-borne pathogens (e.g., *Anaplasma, Babesia* [135, 136]) or additional climatic variables, such as humidity or rainfall. The framework we developed can be enhanced by explicitly accounting for the aforementioned factors, in addition to also explicitly accounting for the impacts of human mobility, animal dispersal animal dispersal (such as deer or migratory birds that can transport ticks across counties) and land cover data. In conclusion, this study presented a scalable, temperature-driven mathematical modeling framework for realistically assessing and predicting the geospatial transmission dynamics and control of Lyme disease in Maryland under current and projected climate change scenarios. Specifically, the study theoretically determined an optimal temperature range (17.0–20.5°C) for maximum abundance of ticks and Lyme disease transmission in the state of Maryland, in addition to predicting the geospatial distribution and burden of the disease under various climate change projections (generally predicting westward shift of Lyme disease under global warming) and highlighted immature ticks and rodent pathways as transmission drivers. Our results also showed that an integrated hybrid intervention (which combines habitat clearance, rodents baiting, and personal protection against exposure to ticks) could lead to Lyme disease elimination in Maryland under moderate (attainable) levels of efficacy and coverage. The methodologies and results developed in this study can be adapted and used to study the impact of climate variables on the geospatial dynamics of Lyme disease in other geographies or regions, as well as for other vector-borne diseases in general.

## Data Availability

All data used/produced in the present work are contained in the manuscript

## CRediT authorship contribution statement

**SSM**: Conceptualization, Data curation, Formal analysis, Methodology, Software, Validation, Visualization, Writing – original draft, Writing – review & editing. **ABG**: Conceptualization, Funding acquisition, Methodology, Project administration, Supervision, Validation, Writing – original draft, Writing – review & editing.

## Acknowledgments

ABG acknowledges the support, in part, of the National Science Foundation (Grant Number: DMS2052363; transferred to DMS-2330801).

